# AI enabled exome and transcriptome liquid biopsy platform spanning the continuum of care in oncology

**DOI:** 10.1101/2025.05.23.25328255

**Authors:** Jim Abraham, Valeriy Domenyuk, Maria Perdigones Borderias, Sergey Klimov, Sourabh Antani, Takayuki Yoshino, Elisabeth Heath, Emil Lou, Stephen Liu, John Marshall, Wafik El-Deiry, Anthony Shields, Martin Dietrich, Yoshiaki Nakamura, Takao Fujisawa, George Demetri, Anna Barker, Joanne Xiu, Dominic Sacchetti, Seth Stahl, Rob Hahn-Lowry, Adam Stark, Jeff Swensen, George Poste, David Halbert, Matthew Oberley, Milan Radovich, George Sledge, David Spetzler

## Abstract

**Background:** Effective clinical management of patients with cancer requires highly accurate diagnosis, precise therapy selection, and highly sensitive monitoring of disease burden. Caris Assure is a multifunctional blood-based assay that couples whole exome and whole transcriptome sequencing on plasma and leukocytes with advanced machine learning techniques to satisfy all three clinical testing needs on one platform.

**Patients and Methods:** Caris Assure for therapy selection was CLIA validated using 1,910 samples. 376,197 tissue profiles along with 7,061 paired blood and tissue profiles were used to engineer features for three machine learning models. The MCED model was trained on 1,013 patients and validated on an independent set of 2,675 patients. The tissue of origin for MCED model was trained on 1,166 samples and validated using 5-fold cross validation. The MRD & Monitoring model was trained on 3,439 patients and validated on two independent sets of 86 patients for MRD and 101 patients for monitoring.

**Results:** For early detection, sensitivities for stages I-IV cancers (n= 284, 129, 90, 23 respectively) were 83.1%, 86.0%, 84.4%, and 95.7%, all at 99.6% specificity (n=2149). The diagnostic first-line procedure for tissue of origin was determined for 8 categories with a top-3 accuracy of 85% for stage I and II cancers. Detection of driver mutations for therapy selection from blood collected within 30 days of matched tumor tissue, demonstrated high concordance (PPA of 93.8%, PPV of 96.8%) using CHIP subtraction. For MRD and recurrence monitoring, the disease-free survival of patients whose cancers were predicted to have an event was significantly shorter than those predicted not to have an event using a tumor naïve approach (HR=33.4, p<0.005, HR=4.39, p=.008, respectively).

**Conclusion(s):** The data presented here demonstrate a unified liquid biopsy platform that uses blood-based whole-exome and transcriptome sequencing coupled with artificial intelligence to address the important clinical needs in multi-cancer early detection, monitoring of MRD and recurrent cancers, and precision selection of molecularly targeted therapies.

## Introduction

Next Generation Sequencing (NGS) technology applied to nucleic acids is now routinely utilized across the continuum of cancer care to measure SNV/INDEL, microsatellite instability status (MSI), tumor mutational burden (TMB), copy number, expression, and fusions^1,2^. Current guidelines recommend concurrent testing of both tissue and blood in multiple tumor types^3,4^. For late-stage disease, NGS of both limited gene panels and whole exomes (comprehensive genomic profiling, CGP) are standard-of-care options in therapy selection for many cancer types ^3,4^. Caris has performed tissue-based clinical whole exome sequencing (WES) and whole transcriptome sequencing (WTS) for more than 350,000 late-stage solid tumor cancer patients, resulting in one of the largest and most comprehensive clinico-genomic data sets in the world^5–7^.

For CGP, tumor tissue has been the gold standard, but recent plasma-based “liquid biopsy” tests have attempted to detect tumor-specific genetic alterations without the need for an invasive biopsy^8^. However, most currently available blood-based cancer genomic assays do not account for alterations that reside in the hematopoietic compartment, which can lead to false positives with erroneous therapeutic consequences^9,10^. For early stage disease, detection of minimal residual disease (MRD) via NGS after surgical intervention is increasingly being utilized for some cancers, enabling physicians to identify patients whose disease is at a higher risk of recurrence or could recur more rapidly than others^11,12^. This enables improved clinical trial stratification, for example, to explore more aggressive treatments sooner than might otherwise be standard. It also allows physicians to choose other standard treatment approaches based on the totality of the evidence^13,14^. For the screening and early detection of healthy or high-risk populations, Multicancer Early Detection (MCED) NGS tests are in development to detect the presence of cancer in previously undiagnosed patients and indicate the likely tumor tissue-of-origin. Current MCED tests have low sensitivity for early-stage cancers which limits their utility^15–18^.

The Caris Assure assay is a clinically validated WES/WTS CGP assay optimized for plasma circulating tumor DNA/RNA (a subset of the pool of circulating free total nucleic acids, cfTNA) that can be further utilized to address additional features to increase patient benefit. WES and WTS data were utilized from over 350,000 tumor specimens to select regions for enhanced depth to maximize information relevant to cancer. The nucleic acids from both the plasma and cellular elements in the buffy coat are sequenced independently at high depths; therefore alterations derived from clonal hematopoiesis can be correctly distinguished from tumor-derived alterations^9^. The blood-based assay’s performance has been validated in detecting SNVs, INDELs, structural variants, CNA, TMB, and MSI against the matched tissue-based test to achieve high sensitivity and high PPV relative to the tissue as the gold standard. These features are essential in guiding therapy for patients with advanced stage disease and those with disease recurrence.

The Caris Assure assay was coupled with bespoke Assure Blood-based Cancer Detection AI (ABCDai) machine learning models to provide sensitive and specific signals that address testing requirements across the spectrum of cancer care from early detection, diagnosis, and therapy selection to MRD and monitoring.

## Methods

### Patient Samples

Samples were acquired either through the Caris Biorepository, Discovery Life Sciences, Indivumed, or through clinical testing from over 80 different institutions. Early-stage cancer samples were acquired prior to treatment for MCED studies, while some later stage patient samples may have been post-treatment. Specific treatment data are unknown. Studies were conducted under IRB approval WCG: TCBIO-001-0710. This study was conducted in accordance with the guidelines of the Declaration of Helsinki, Belmont report, and U.S. Common rule. In keeping with 45 CFR 46.101(b)(4), this study utilized retrospective, de-identified clinical data. Therefore, this study was considered IRB exempt and no patient consent was necessary from the subject. Information regarding consent and the source of all samples used in these studies can be found in Supplementary Table 1.

Disease free survival was calculated as the time from surgery to either relapse/death or censoring. Samples were excluded from the analysis if there were incomplete clinical follow-up data for calculation of survival, data entry issues, surgery after first blood draw, relapse before blood draw, CNS malignancy, and cases that were from patients with metastatic disease or not treated in the adjuvant curative setting. Analysis was restricted to stage II and III cancers and was performed for a 3-year time period wherein patients were right-censored at 3 years if they had a longer follow-up (regardless of event status past 3 years).

### Assure Workflow

The Caris Assure workflow prepares libraries and sequences both cfDNA and cfRNA simultaneously in a single sequencing run using hybridization/capture methodology. Specific regions were targeted using a baited captured pull-down of DNA and cDNA (RNA) exonic regions for subsequent library preparation, with subsequent deconvolution of the DNA and RNA sequencing reads bioinformatically. Thus, Caris Assure provides WES and WTS data from the cfTNA specimen. The cfTNA was extracted from plasma using a novel, high-throughput automated method, customized from the DSP Virus/Pathogen Midi kit (Qiagen, custom) and Hamilton Star liquid handler system. First plasma is mixed with standard lysis buffer with high concentration of guanidinium salts, dithiothreitol (DTT), and carrier RNA to inhibit RNAases. Second, proteinase K is added to degrade protein RNAase molecules. Third, SDS is added to standard binding buffer to lyse circulating microvesicles which were protecting RNA. To perform simultaneous DNA and RNA sequencing, custom cDNA primers were used (IDT, Coralville, IA, USA; GeneLink, Orlando, FL, USA). RNA was chemically tagged during first strand cDNA synthesis. Custom hybrid pull-down panels of baits were designed to enrich for 720 clinically relevant genes at high coverage and high read-depth and an additional >20,000 genes at lower depth. Sequencing was performed on a NovaSeq 6000 instrument (Illumina, San Diego, CA, USA) using recommended reagents. Sequencing data was extracted into split FASTQ files (RNA and DNA) for further processing using Caris’s bioinformatics pipeline. For RNA, Spliced Transcripts Alignment to a Reference (STAR) software was used for alignment, trimming, and fusion detection using the RNA FASTQ files from TNA split pipeline. Transcripts per million (TPM) molecules were generated using the Salmon expression pipeline. All paired buffy coat samples had gDNA extracted using either the automated DSP DNA Midi kit (Qiagen, Cat# 937255) or the Mag-Bind Blood & Tissue DNA HDQ 96 Kit (Omega Biotek, Cat# M6399-01). Sequencing libraries were prepared using HyperPrep kits, HyperPure Beads, custom primer mixes and baits, (KAPA/Roche) and custom cDNA primers (IDT/GeneLink, AeG2A). Sequencing was performed on a NovaSeq System with NovaSeq 6000 S4 Reagent Kits (Illumina). Separate sequencing was performed on the nucleic acids extracted from the matched buffy coat to identify potential germline variants and variants arising from clonal hematopoiesis. Likely CHIP variants in plasma were subtracted from the set of pathogenic and likely pathogenic variants carried into subsequent analysis and reporting. All samples passed CLIA QC requirements.

### Machine Learning Methodologies for ABCDai base models

ABCDai models are gradient-boosted decision trees built with XGBoost^19^. The parameters used are 500 estimators, no maximum depth, 0.75 subsample, 0.75 colsample_bytree and 42 as the random seed. These parameters were selected heuristically based on our previous studies of separate liquid biopsy data to optimize performance and generalization. Each ABCDai model is built in two phases. The first phase is used for feature selection and generates XGBoost models for each of the nine foundational feature sets(pillars): Fusionome, Mutationome, Motifome, Fragmentome, Copyome, Entropyome, PositionomeNU, PositionomeTF and Transcriptome. The second phase extracts feature importances from each of the phase 1 models and creates a panomic feature set using the top 500 features from each pillar-based model. To address the needs of MCED, diagnostic pathway prediction, and for MRD/Monitoring, three ABCDai models were constructed in this study. Where applicable, stratified flow cell-grouped k-fold cross-validation was employed to mitigate flow cell bias while maintaining balanced label proportions.

### Fusionome for ABCDai

The BAM file was analyzed for reads with clips of 12 or more bases. The clips are analyzed for potential secondary alignments. Reads aligned to chrX and chrY were excluded from analysis.

### Mutationome for ABCDai

The BAM file for the sample was analyzed for the presence of SNV/Indel mutations using Mutect2. From the variants detected, those with a predicted effect of Frameshift, Missense, Insertion, Deletion or Nonsense were selected. For each SNV, a ref-context was created with a reference sequence from one base to each side of the variant position, and an alt-context was created by replacing the reference allele with the variant allele in the 2^nd^ position of the ref-context. The number of variants per ref-context and alt-context are normalized by the total count of variants across all the ref-contexts and alt-contexts respectively.

Variant-supporting reads for pathogenic and likely pathogenic somatic variants with high mapping and base quality were quantified across the whole exome and normalized per million aligned on-target reads. This calculation was referred to as tumor parts per million (PPM) and was used as a measure of assessing tumor fraction in the plasma sample.

### Motifome for ABCDai

Each read in the BAM file is analyzed, and those that are measured as duplicates, secondary alignments, or have a mapping quality under 30 are filtered out. From the remaining reads, the left and right 4-mer motifs (first and last four bases of the read sequence) were extracted after accounting for read orientation. The proportion of each of the 256 unique motifs was then stored as a 256-length vector. Finally, the normalized shannon entropy was computed from the 256-length motif vector set and stored as an additional feature^20,21^

### Fragmentome for ABCDai

The sample BAM file was then analyzed and the reads with read lengths of ≥50 bases and <250 bases were selected. These reads were analyzed for the presence of smaller fragments.

### Copyome for ABCDai

A CNVkit reference was built from 8 independent normal plasma samples. CNVKit was run in hybrid sequencing method using HMM segmentation. A segment weight cutoff of 50 was used.

### Entropyome for ABCDai

The BAM file was analyzed and for each contiguously baited region with 10 or more reads with mapping quality of 30 or higher, the following features were generated: ratio of number of unique alignment start positions to number of reads, ratio of number of unique fragment lengths to number of reads, entropy of the unique start positions for reads between 80 and 250 bases long, and unique fragment lengths for reads between 80 and 250 bases long.

### Positionome NU & Positionome TF for ABCDai

The BAM file was analyzed, and for each gene, reads with the midpoint of alignment (the position equidistant from the alignment start and end) between the start and end of the first exon were selected. These reads were further filtered into reads with lengths between 146 and 170 (called single-nucleosomes(NU)) and reads with lengths between 60 and 100 (called Transcription Factor(TF)-like). For each gene, the following features were extracted: ratio of count of single-nucleosome reads before exon 1 and count of single-nucleosome reads within exon 1, ratio of count of TF-like reads before exon 1 and count of TF-like reads within exon 1, and total number of reads of both types between the start and end of exon 1. To be included for analysis a minimum of 150 total filtered reads must be present within and before exon 1.

### Transcriptome for ABCDai

RNA read counts are normalized to trimmed mean of M-values (TMM). An expression-reference for the TMM calculation was created using mean expression values from 8 normal samples. To create the heatmaps using the transcriptome data, we first applied a z-transformation to the values of the genes chosen for the MCED model followed by hierarchical clustering on both the samples and the gene features.

## Results

### Assure Cell-free Nucleic Acid and gDNA Concordance to Tissue

To assess accuracy, 166 de-identified matched plasma/tissue specimens were analyzed retrospectively across various cancer types and the Caris Assure assay was compared to the Caris MI Exome/Transcriptome FFPE tissue assay. Plasma and buffy sequencing metrics can be found in the Supplementary Materials.

Detection of driver mutations in metastatic cancer, where blood was collected within 30 days of matched tumor tissue, demonstrated high concordance with a PPA of 93.8% and PPV of 96.8% and was either comparable or superior to other cfDNA assays^22,23^ (Figure 1A). CHIP correction proved to be essential as over 50% percent of patients had CHIP mutations, including KRAS, ATM, and CHEK2. If only plasma is analyzed, these mutations could be interpreted as tumor-derived, leading to improper therapy selection. A detailed summary of Assure validation results are shared in the Supplementary Materials.

**Figure 1.**
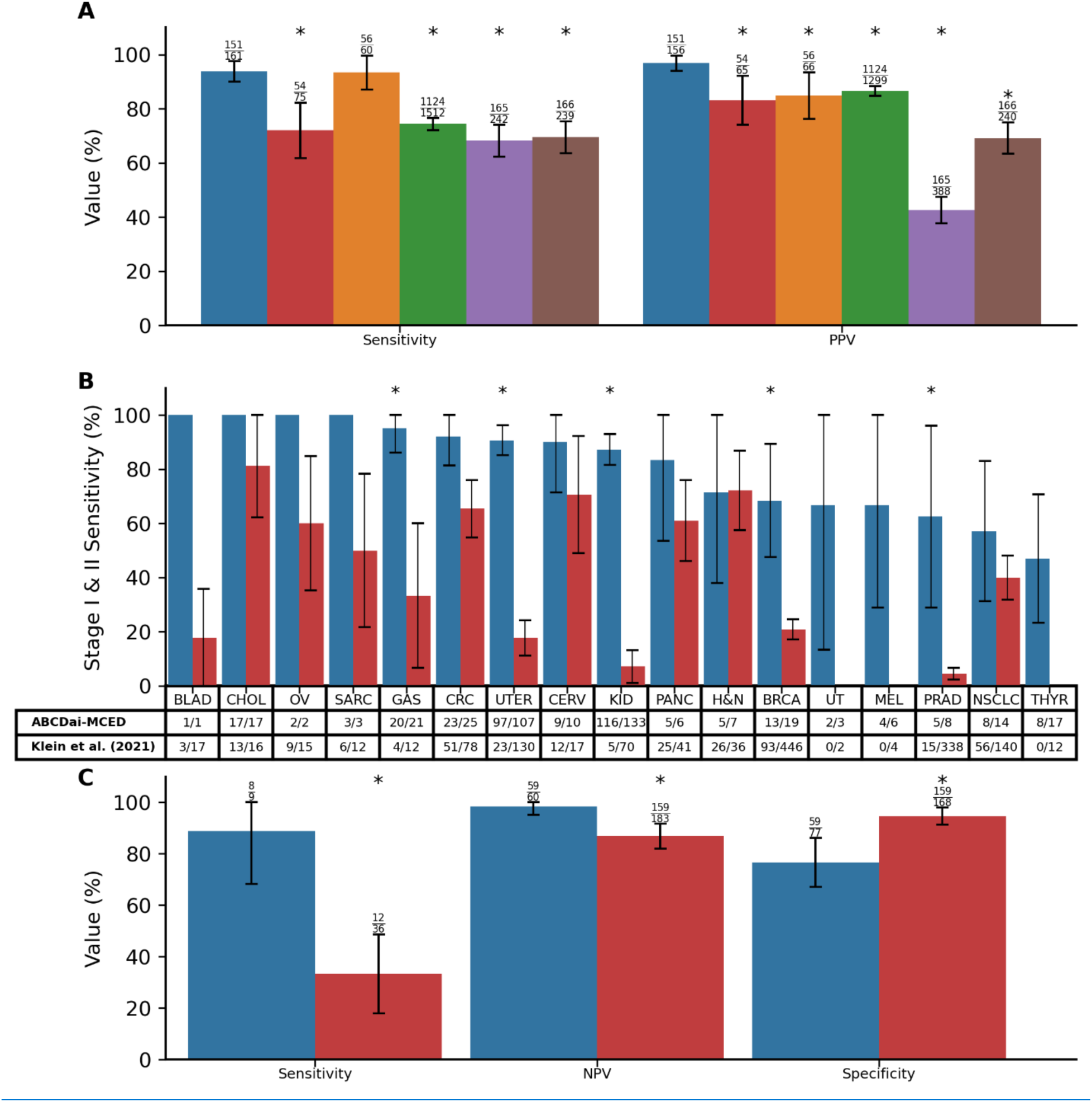
Assure and ABCDai Benchmarking. A) Comparison of Caris Assure performance with published results of plasma-based genotyping^22,23^. The error bars indicate the 95% CIs and the asterisks (*) signify a significant difference of the TP/FN (for sensitivity) or TP/FP (for specificity) proportions between tests. Blue, Caris Assure; Red, Aggarwal; et al. (2018); Orange, Leighl et al. (2019); Green, Iams et al. (2024); Purple, Finkle et al. (2021); Brown, Bayle et al. (2024). B) Comparison of performance of ABCDai-MCED in Stage I and Stage II cancers against the published performance of a commercially available early detection product^51^. The error bars indicate the 95% CIs and the asterisks (*) signify a significant difference of the TP/FN proportions between tests. Blue, ABCDai-MCED; Red, Klein et al. (2021). C) Comparison of sensitivity, NPV, and specificity of ABCDai-M&M and a published tumor information naïve MRD product^52^. The error bars indicate the 95% CIs. Blue, ABCDai-M&M; Red, Nakamura et al. (2024). Abbreviations: BLAD, Bladder; CHOL, Cholangiocarcinoma; OV, Ovarian; SARC, Sarcoma; GAS, Gastric; CRC, Colorectal Cancer; UTER, Uterus; CERV, Cervical; KID, Kidney; PANC, Pancreatic; H&N, Head and Neck; BRCA, Breast Cancer; UT, Urinary Tract; MEL, Melanoma; PRAD, Prostate Adenocarcinoma; NSCLC, Non-small-cell Lung Cancer; THYR, Thyroid.

### Assure Blood-based Cancer Detection AI (ABCDai) Algorithm Development

ABCDai models are generated using nine sets of features (pillars): Copyome, Fusionome, Mutationome, Transcriptome, Entropyome, PositionomeNU, PositionomeTF, Motifome, and Fragmentome (Figure 2). The first four of these pillars are traditionally studied in tissue-based cancer research (features encompassing copy number variations, fusions, mutations, and gene expression, respectively) and features for these were engineered using 376,197 tissue cases. The remaining five are unique to the liquid biopsy space and features for them were engineered using the 7,061 matched tissue and plasma cases.

**Figure 2.**
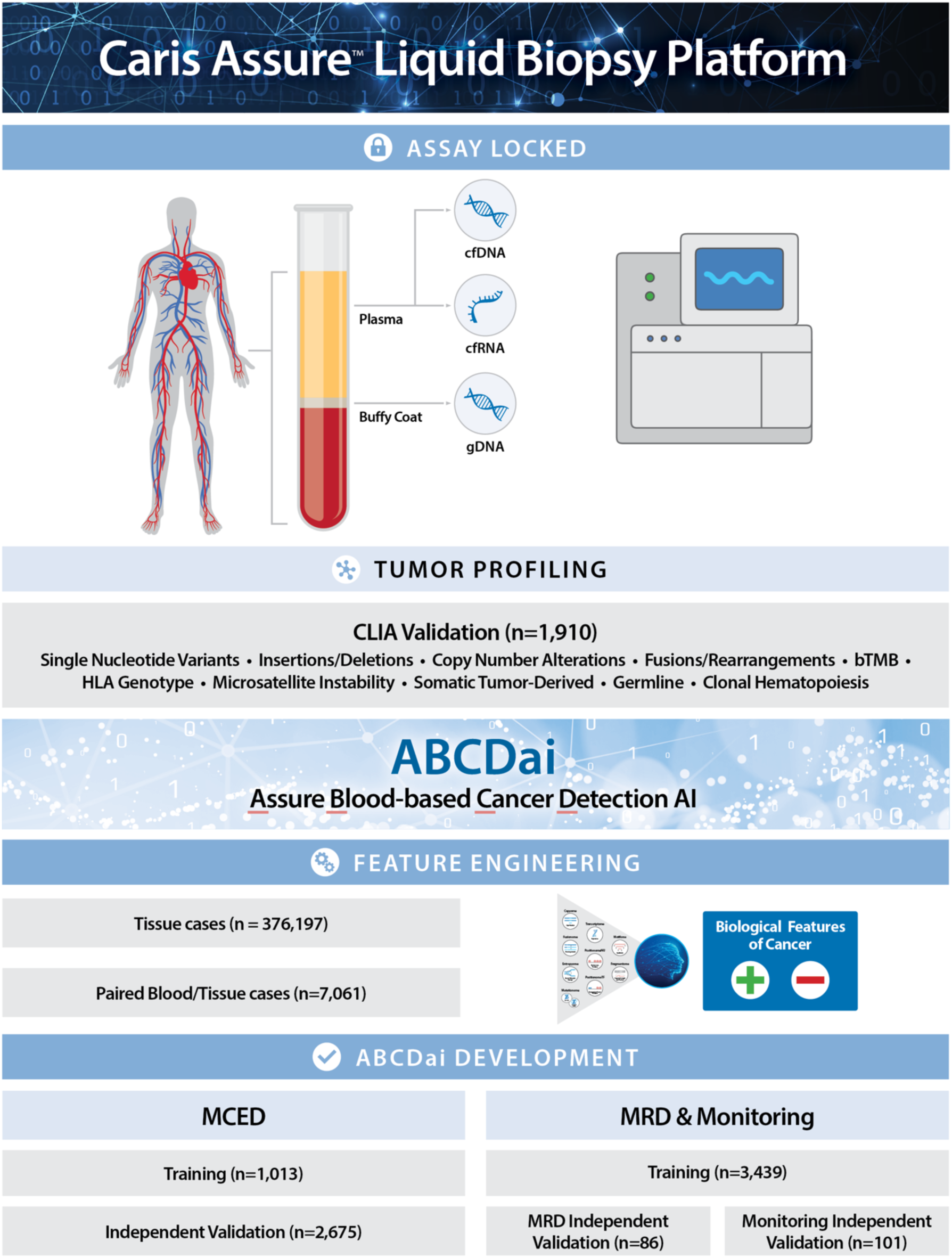
Assure workflow and Assure Blood-based Cancer Detection AI development and validation. DNA and RNA are extracted from the subject’s plasma along with gDNA from the subject’s buffy coat. These analytes are then sequenced and processed through bioinformatic workflows to generate biologically relevant feature sets (Pillars). The pillars are fed into a panomic machine learning algorithm (ABCDai) to generate a prediction. The diagram also describes assay and AI development with samples used in each study. ABCDai-GPS was trained using samples from 660 patients with stage I and II cancers and 506 normal donors. Performance for ABCDai-GPS was evaluated using 5-fold cross validation. All data is stored in deidentified Caris proprietary database and may be requested per Data Availability statement.

The single molecule information obtained from the plasma stems from the specific properties of cfDNA in circulation. Although multiple sources account for cfDNA in the plasma^24^, the most common origin is apoptosis. Apoptosis leads to DNA breaks by caspase-activated DNase and in turn releases DNA fragments wrapped around nucleosomes, with peaks of 167 base pairs, consistent with the apoptotic ladder and the size of a nucleosome and its linker protein. The contents of these dying cells can eventually enter the blood stream and be detected if the rate of clearance is insufficient, such as during cancer^25^.

This non-random fragmentation of DNA ‘protected’ by nucleosomes or other proteins, such as transcription factors^18^ inherently underlies important cellular footprints due to the presence of nucleosomes at those positions. Nucleosome organization, positioning, spacing, and variability partially regulate transcriptional activity ^26^. Furthermore, the transcriptional process induces cycles of nucleosome eviction and reoccupation^23^ which can be observed deep into the gene body ^27^. These positional nucleosome (and transcription factor) patterns are thus distinct depending on the tissue that releases them and can be used for cancer detection, tissue of origin prediction^18^and as proxies for gene expression in these distant cells^28,29^.

The DNA wrapped around the nucleosome also has plasma specific characteristics that stem from variations in its cleavage. Nucleosomes in normal non-somatic cells are more methylated, which causes tighter binding and, in-turn, leads to consistent cleavage between nucleosomes, resulting in frequent ^20,26,27^167 base-pairs^1,24,30^ fragments and, less frequently, multiples of that size reflecting di or tri-nucleosome packing. Conversely, cancer cells often have hypomethylated nucleosomes, which leads to looser binding and opens alternative cleavage sites, resulting in a shift in the distribution of fragment size towards a shorter length. This can be further compounded by nuclease type variations (and thus their preferential cleavage sites), their relative expression, DNase hypersensitive site accessibility, and nucleosome positioning^31^. Consequently, these additional cleavage sites lead to variations in cfDNA fragment ends^24^.

The first of the five feature pillars which aim to capture these biological phenomena is the Entropyome which captures the fragment diversity, due to nucleosome displacement, inside gene/exons as increased diversity was shown to correlate to gene expression^27,32^. The second is the PositionomeNU (nucleosome position) which captures the relative prevalence of nucleosome sized fragments upstream of exon 1 to measure variations near the transcription start site^18^. The third is the PositionomeTF (transcription factor position) which captures the relative prevalence of transcription factor sized fragments upstream of exon 1 to measure variations near the transcription start site^18^. The fourth is the Fragmentome which captures fragment length distribution changes across the exome. Finally, the fifth is the Motifome which captures patterns in nucleotide end motifs^20^.

To evaluate the capabilities of these feature pillars in measuring cancer, we analyzed a few individual exemplar features known to be aberrant from tissue studies. Copyome analysis revealed significant recurrent chromosomal alterations across multiple cancer types. There was a significantly (h=300, p=4.e-57, eta^2^=0.08) elevated level of Copyome derived aneuploidy in 8 high-stage cancer types (detailed in Supplementary Table 2) with at least 30 samples each (Figure 3A) with only ovarian (p=0.08) cancer not reaching post-hoc significance against normal samples. In high-stage NSCLC (Figure 3B), we observed a deletion at 3p, an amplification at 3q26-29, and a deletion at 6q24-25^33^. For high-stage colorectal cancer (Figure 3C), notable findings include an amplification at 7p, a deletion at 8p, and an amplification at 13q ^34,35^. Both high-stage breast cancer (Figure 3D) and prostate cancer (Figure 3E) exhibited an amplification at 8q and deletions at 8p, 13q, and 16q, with breast cancer additionally showing a deletion at 17p ^36,37^. A hierarchical clustering analysis of the top 500 expression features used in the creation of the ABCDai-MCED model shows transcriptional patterns associated with normal and cancer samples (Figure 4A). At a gene level, we see significant (ANOVA P<0.05) differences in expression across a subset of genes found as part of the COSMIC Cancer Gene Census^38^. This includes genes with evidence of some tumor suppression ability such as KDM5C, NF2^39^ CYLD^40^, and SPOP^41^ (Figure 4B) and the genes showing evidence of overexpression in cancers such as BMPR1A^42^, GMPS^43^, POLQ^44^, PSIP1^45^, and FLNA^46^ (Figure 4C). The Fragmentome feature, ‘Median Fragment Length’, shows a consistent decrease across the genome for cancer samples (exemplar in Figure 4D). Furthermore, we observed 75% (193/256) of end motifs (Figure 4E), with a significant difference between cancer and non-cancer patients. PositionomeNU showed significantly (ANOVA p<0.005) higher levels of nucleosomes before exon 1 for tumor suppressor genes PMS2^34^ and NF2^39,47^ (Figure 4F). Conversely, we see sparser nucleosome occupation for the oncogenes MYCN ^48^ and TERT^49^(Figure 4G).

**Figure 3.**
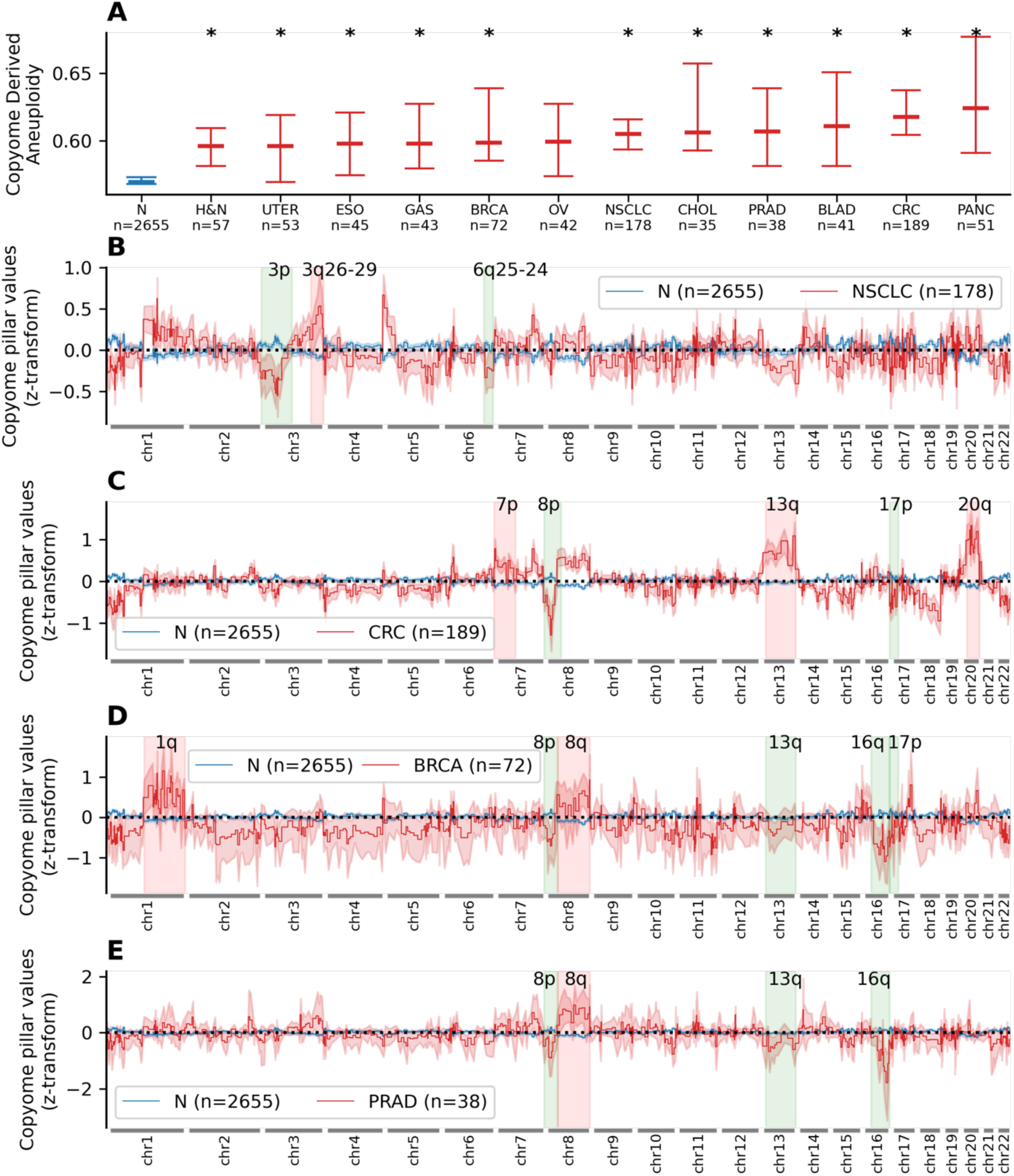
Copy Number Comparison. A) Median Copyome derived aneuploidy with 99% CI for normal and late-stage cancer samples. Copyome derived aneuploidy was computed as the fraction of Copyome bins where Copyome pillar value was < -0.1 or > 0.1. Asterisks (*) indicate significance in Copyome aneuploidy between specific cancer samples and normal. B) Mean z-transformed Copyome pillar values for 2655 normal samples vs 178 late-stage NSCLC samples with 95% CI error band. The highlighted regions show the cytobands known to harbor higher number of CNV variants in NSCLC. The 2 sample ks-test with mean Copyome value for the given band between normal and NSCLC samples showed a p-value of 3.33e-16 (KS test statistic: 0.39) for 3p, 5.37e-9 (KS test statistic: 0.24) for 3q26-29, and 6.68e-7 (KS test statistic: 0.21) for 6q25-24. C) Mean z-transformed Copyome pillar values for 2655 normal samples vs 189 late-stage CRC samples with 95% CI error band. The highlighted regions show the cytobands known to harbor higher number of CNV variants in CRC. The 2 sample ks-test with mean Copyome value for the given band between normal and CRC samples showed p-value of 2.89e-15 (KS test statistic: 0.31) for 7p, 7.77e-16 (KS test statistic: 0.33) for 8p, 7.77e-16 (KS test statistic: 0.44) for 13q, 7.77e-16 (KS test statistic: 0.42) for 17p, and 7.77e-16 (KS test statistic: 0.45) for 20q. D) Mean z-transformed Copyome pillar values for 2655 normal samples vs 72 late stage breast cancer samples with 95% CI error band. The highlighted regions show the cytobands known to harbor higher number of CNV variants in breast cancer. The 2 sample ks-test with mean Copyome value for the given band between normal and breast cancer samples showed p-value 2.82e-9 (KS test statistic: 0.37) for 1q, 9.26e-7 (KS test statistic: 0.32) for 8p, 4.11e-6 (KS test statistic: 0.30) for 8q, 3.45e-5 (KS test statistic: 0.28) for 13q, 1.4e-14 (KS test statistic: 0.47) for 16q, and 2.33e-9 (KS test statistic: 0.38) for 17p. E) Mean z-transformed Copyome pillar values for 2655 normal samples vs 38 late stage prostate cancer samples with 95% CI error band. The highlighted regions show the cytobands known to harbor higher number of CNV variants in prostate cancer. The 2 sample ks-test with mean Copyome value for the given band between normal and prostate cancer samples showed p-value 3.74e-3 (KS test statistic: 0.28) for 8p, 3.66e-5 (KS test statistic: 0.37) for 8q, 3.16e-4 (KS test statistic: 0.33) for 13q, 1.91e-11 (KS test statistic: 0.56) for 16q. High Grade Sample N numbers can be found in Supplementary Table 2. Abbreviations: BLAD, Bladder; BRCA, Breast Cancer; CHOL, Cholangiocarcinoma; CRC, Colorectal Cancer; ESO, Esophageal; GAS, Gastric; H&N, Head and Neck; NSCLC, Non-small-cell Lung Cancer; N, Normal; OV, Ovarian; PANC, Pancreatic; PRAD, Prostate Adenocarcinoma; UTER, Uterus.

**Figure 4.**
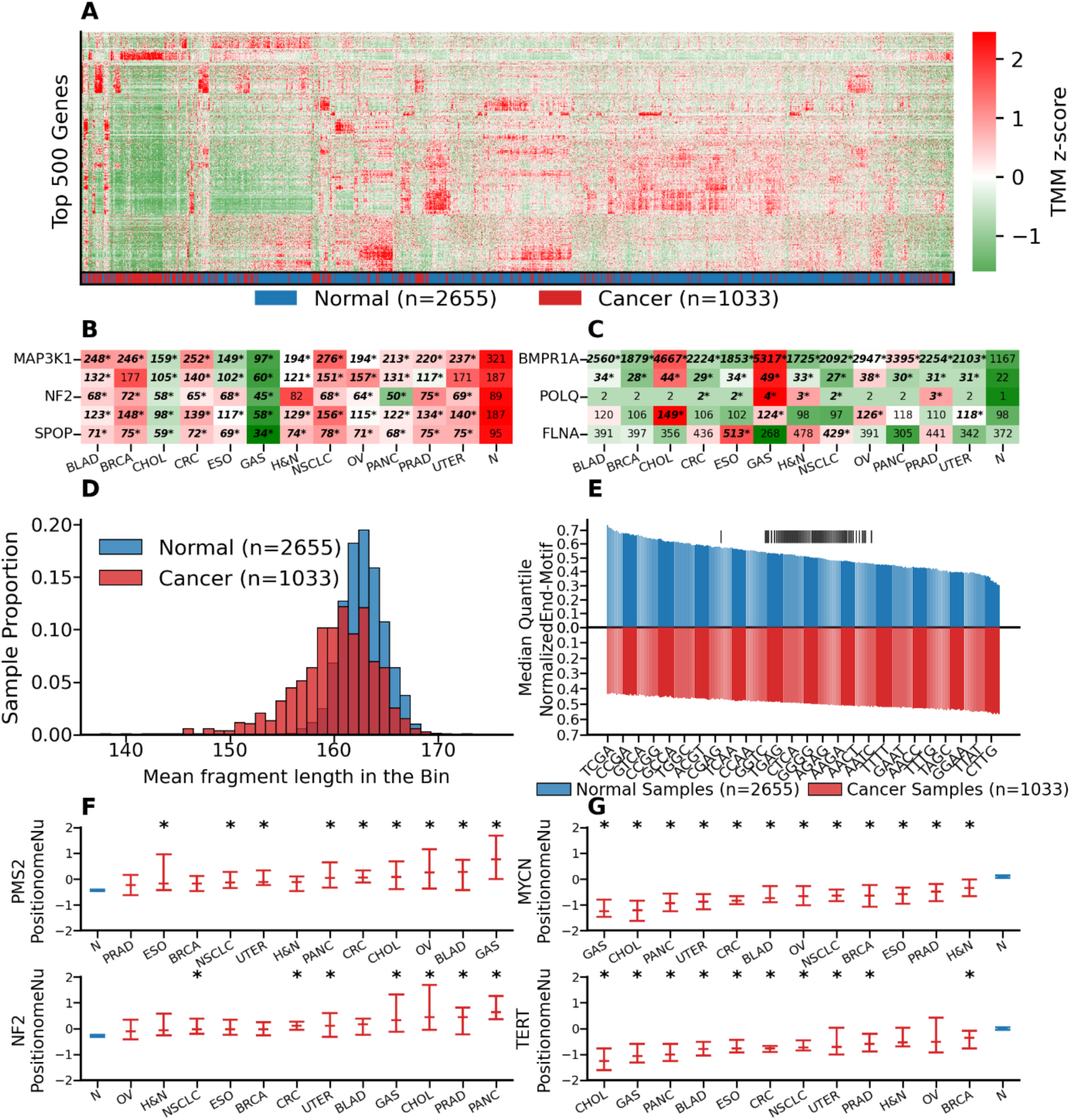
Exemplar Single Molecule and Transcriptome Features used in ABCDai. A) Hierarchical clustered heatmap of z-score normalized expression values for late-stage cancer (III/IV) and normal samples using the 500 genes selected in the independent MCED model. B) TMM values for a set of genes having reduced expression in cancer samples (MAP3K1 h=463, p=1e-91, eta^2^=0.12; KDM5C h=444, p=1e-87, eta^2^=0.12; NF2 h=329, p=3e-63, eta^2^=0.09; CYLD h=428, p=4e-84, eta^2^=0.12; SPOP h=347, p=5e-67, eta^2^=0.10). An asterisk (*) represents a significant difference for specific cancer TMM values vs. Normal TMM values. C) TMM values for a set of genes having overexpression in cancer samples (BMPR1A h=441, p=4e-87, eta^2^=0.12; GMPS h=308, p=6e-59, eta^2^=0.09; POLQ h=111, p=2e-8, eta^2^=0.05; PSIP1 h=96, p=3e-15, eta^2^=0.02; FLNA h=72, p=1e-10, eta^2^=0.02). An asterisk (*) represents a significant difference for specific cancer TMM values vs. Normals. D) Mean fragment length across an approximate 5MB bin for normal samples (blue) (mean=162.8, std=2.1) are significantly shorter (t=-27, p=1e-147) than late-stage cancer samples (red) (mean=160.0, std=4.1). E) Median quantile normalized motif proportions between Normal (blue) and late-stage Cancer (red) sorted by magnitude of difference. A vertical line above a motif (|) indicates a non-significant (p>0.05) 2-sample independent t-test difference after multiple hypothesis correction. F) Z-score PositionomeNU values for tumor suppressor genes shown to have higher relative nucleosome occupancy before exon 1 in cancer samples (PMS2 h=138, p=1e-23, eta^2^=0.15; NF2 h=184, p=5e-33, eta^2^=0.06). An asterisk (*) over a distribution indicates a significant difference for specific cancer Z-score PositionomeNU values vs. Normals G) Z-score PositionomeNU values for oncogenes shown to have lower relative nucleosome occupancy before exon 1 in cancer samples (MYCN h=584, p=1e-117, eta^2^=0.17; TERT h=487, p=1e-96, eta^2^=0.14). An asterisk (*) over a distribution indicates a significant difference for specific cancer Z-score PositionomeNU values vs. Normals. Cancer type visualizations are filtered to those with at least 30 samples. High Grade Sample N numbers can be found in Supplementary Table 2. Abbreviations: N, Normal; BLAD, Bladder; CHOL, Cholangiocarcinoma; OV, Ovarian; GAS, Gastric; CRC, Colorectal Cancer; PANC, Pancreatic; H&N, Head and Neck; BRCA, Breast Cancer; PRAD, Prostate Adenocarcinoma; NSCLC, Non-small-cell Lung Cancer; UTER, Uterus; ESO, Esophageal.

Three ABCDai models were created using these 9 feature pillars to address various clinical needs (Figure 2). ABCDai-MCED was created for the purpose of multi-cancer early detection. ABCDai-GPS (Genomic Probability Score) predicts which diagnostic pathway ^50^ is optimal given a positive ABCDai-MCED result. And finally, ABCDai-M&M was created to detect minimal residual disease and monitor for recurrence across a variety of cancers.

### Validation of ABCDai-MCED for Multi Cancer Early Detection

ABCDai-MCED was trained using the aforementioned features on samples from 1,013 patients (507 patients with cancer and 506 normal donors). A single model was trained on the entirety of these data and performance was assessed on an independent set of 2,675 patients (526 patients with cancer and 2,149 samples from normal donors), with demographics as described in Table 1.

**Table 1.**
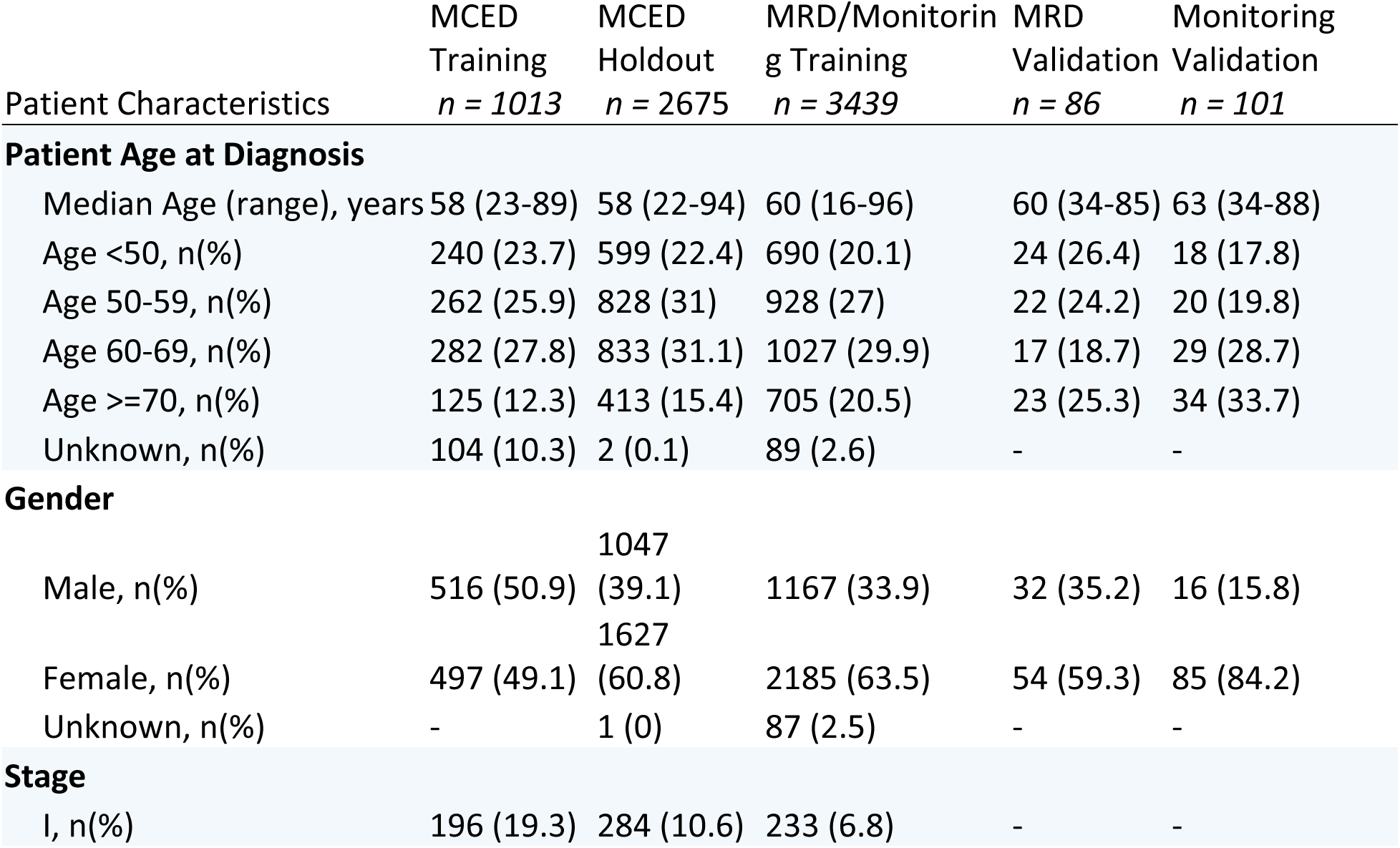
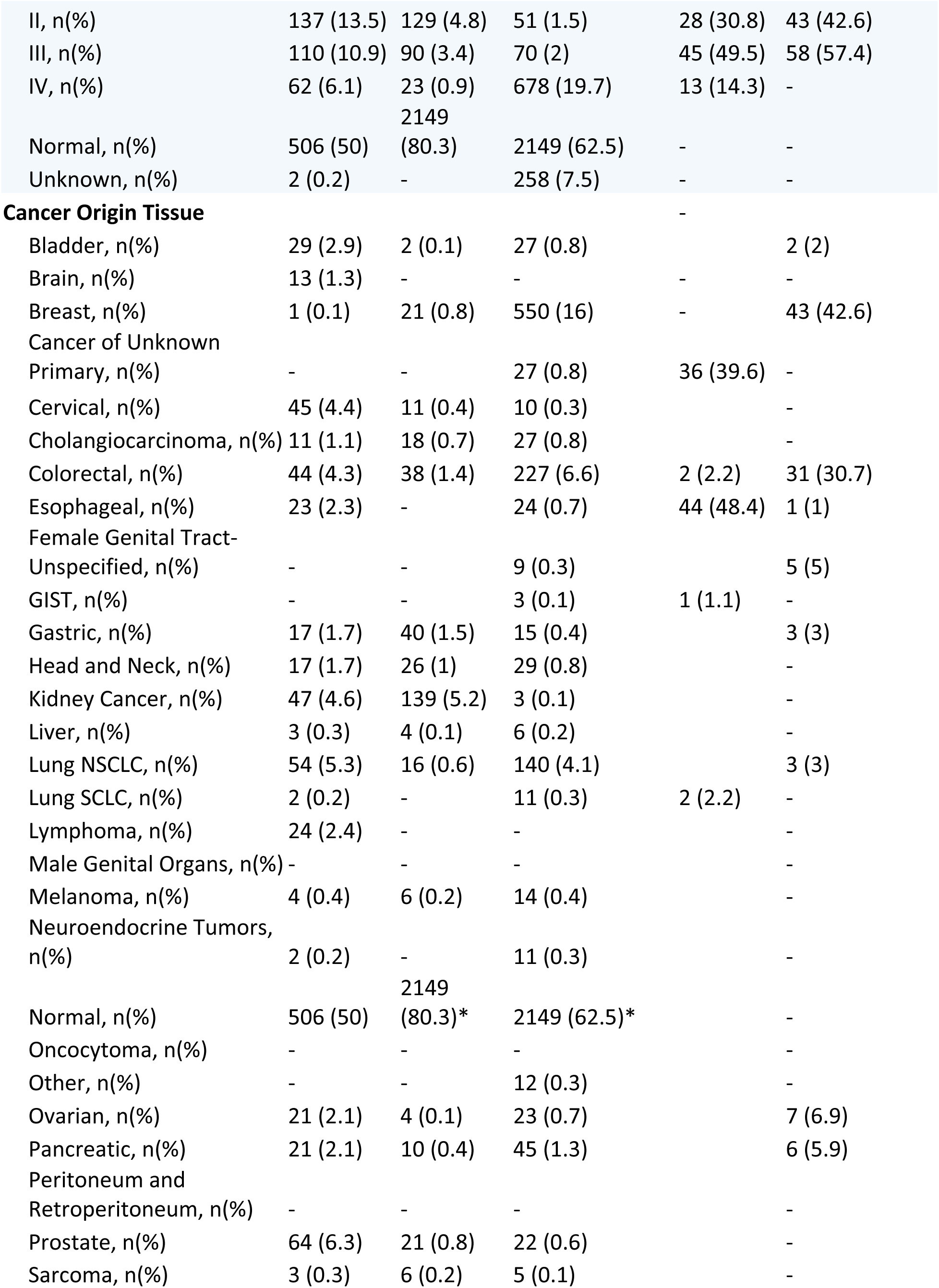
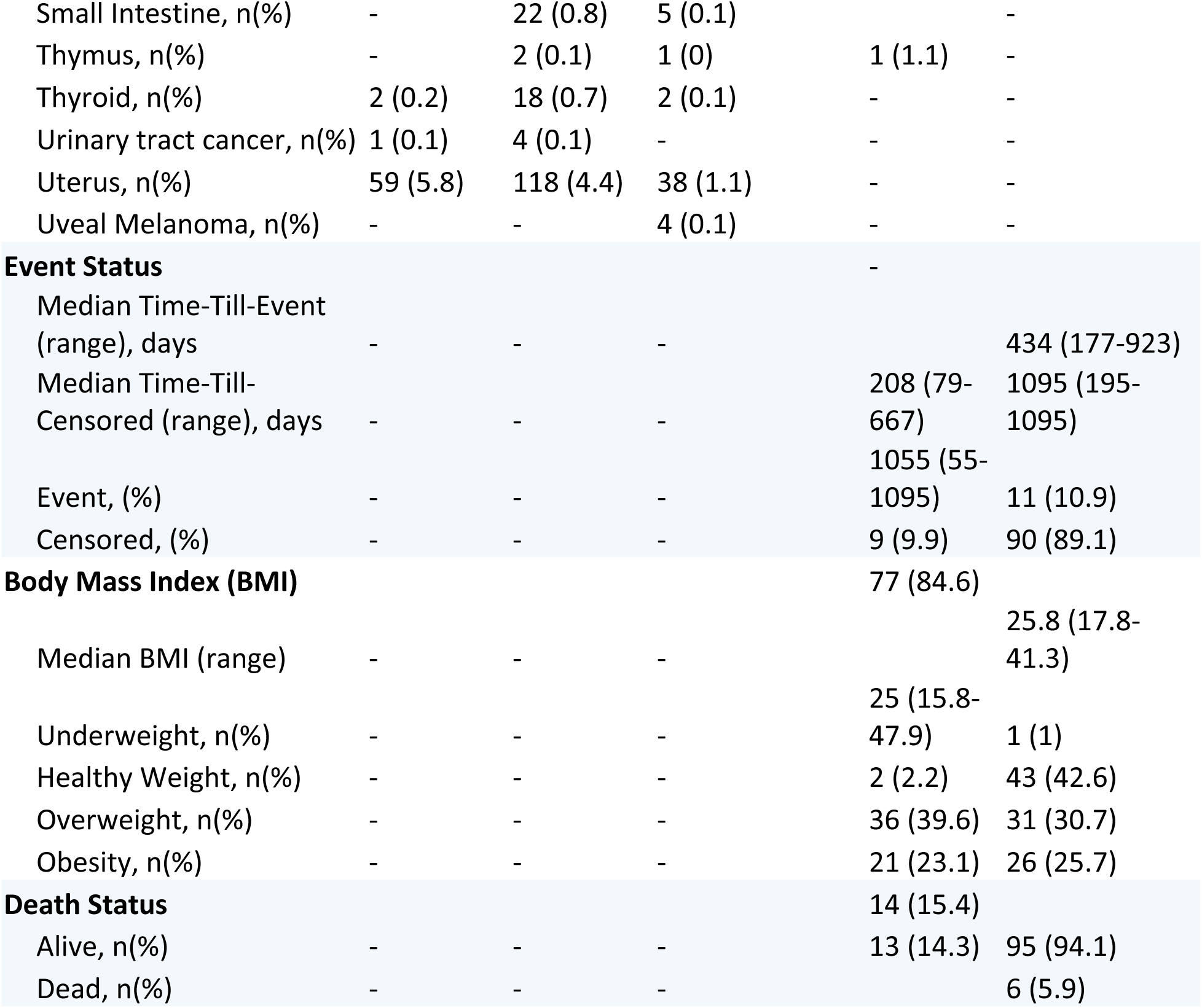
Demographic tables for the MCED, MRD and Monitoring cohorts. MCED samples do not have event status or information related to body mass index. *:Same normal cohort was used for MCED holdout and for MRD/Monitoring training.

ABCDai-MCED’s performance on the stratification of blood samples from the independent cohort of patients with cancer versus those with no reported cancer resulted in sensitivities for stages I-IV (n= 284, 129, 90, and 23, respectively) of 83.1%, 86.0%, 84.4%, and 95.7% at 99.6% specificity (n=2149). Sensitivity by stage and tumor type is shown in Supplementary Figure 1, and detailed metrics can be found in Supplementary Table 3. ABCDai-MCED’s predicted probability, analyzed as a continuous metric, exhibited a general increase with tumor fraction. The median predicted probabilities for samples in the first, second, third, and fourth quartiles of tumor fraction were 0.965, 0.959, 0.983, and 0.996, respectively (Supplementary Figure 2). In a comparative analysis of the performance of ABCDai-MCED with published results of a commercially available MCED product in early-stage cancers, ABCDai-MCED showed increased sensitivity across most cancer types where data was available. ABCDai-MCED showed significantly increased sensitivity in gastric (p<0.001), uterus (p<0.001), kidney (p<0.001), breast (p<0.001) and prostate cancers (p<0.001). Full results are shown in Figure 1B.^51^

### Validation of ABCDai-GPS for Diagnostic Pathway Prediction

Proper intervention requires knowledge of the tissue of origin, and the molecular information provided by Caris Assure can assist in this determination by predicting the diagnostic pathway that can confirm the presence of cancer. Diagnostic pathway prediction is focused on those tumor types that are most common and thus provide the most utility (colonoscopy, an abdominal\chest CT, endoscopy, liver ultrasound, mammograph with MRI, a neck ultrasound, a pelvic ultrasound, and a prostate-specific antigen test). ABCDai-GPS was trained using treatment-naïve samples from 660 patients with stage I and II cancers and 506 normal donors. Performance was evaluated using 5-fold cross validation (Supplementary Table 4). ABCDai-GPS predicted the diagnostic pathway for 100% of the ABCDai-MCED positive calls with a top-3 accuracy of 85% for stage I and II cancers (Supplementary Figure 3).

### Validation of ABCDai-M&M for MRD and Recurrence Monitoring

ABCDai-M&M was trained using samples from 3,439 patients (1,290 patients with cancer and 2,149 normal donors). A single model was trained on the entirety of these data and performance was assessed on two independent validation cohorts: 86 patients in the MRD study and 101 patients in the recurrence monitoring study with demographics and clinical characteristics as described in Table 1 and therapy, blood draw, and event status shown in Supplementary Figure 4 and Supplementary Figure 5.

Upon applying the ABCDai-M&M model to obtain binary classification in the pan cancer MRD independent validation cohort, wherein samples predicted as ‘cancer features present’ indicated the presence of residual disease, and therefore high risk of an event, significant stratification was observed using the default probability threshold of 0.5 (HR=33.40, p<0.005) (Figure 5A). Significance was retained, at a higher HR, after accounting for various clinicopathological covariates such as stage and age (HR=116.56, p=0.013) (Supplementary Table 5). The ABCDai-M&M model was able to capture 89% of MRD recurrence events with 98% NPV and 76% specificity. Stratification results for breast cancer (HR=5.83, p=0.16) and colorectal cancer (100% sensitivity, p=0.117) can be found in Supplementary Figure 6A and Supplementary Figure 6B, respectively. These results match favorably against the published results of a tumor information naïve MRD product ^52^(Figure 1C). The lead time gained by using ABCDai-M&M with a blood draw as opposed to waiting for imaging to detect recurrence is 261 days on average.

**Figure 5.**
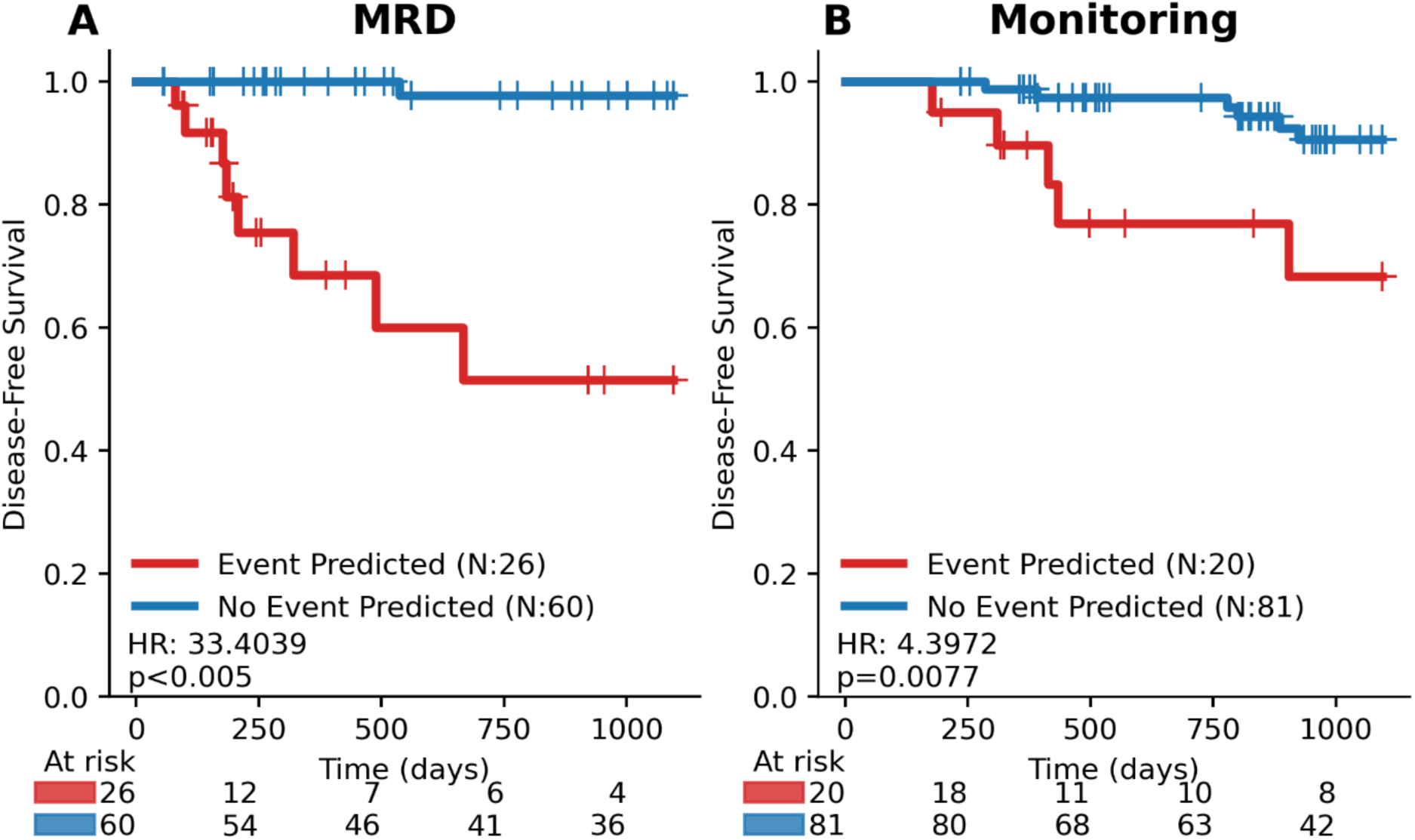
ABCDai-M&M DFS performance on the independent MRD and Monitoring Validation Cohorts. A) Kaplan-Meier survival curves for ABCDai-M&M applied on the independent validation using the default probability threshold of 0.5. B) Kaplan-Meier survival curves for ABCDai-M&M applied on the independent validation using the default probability threshold of 0.5. Univariate hazard ratios are derived from Cox Proportional Hazards models, with significance assessed via log-rank tests. Events are defined as relapse or death, with time measured from the date of surgery for MRD and from therapy initiation for monitoring.

Applied similarly in the recurrence monitoring context, the ABCDai-M&M model demonstrated significant classification ability (HR=4.39, p=0.008) (Figure 5B) towards the pan cancer Monitoring independent validation cohort. Significance was retained, at a higher HR, after accounting for various clinicopathological covariates such as stage and age (HR=6.13, p=0.022) (Supplementary Table 6). The ABCDai-M&M model was able to capture 45% of recurrence monitoring events with 92.6% NPV and 83% specificity. Notably, many recurrence events happen more than 2 years after the last plasma timepoint. Further, the model was able to identify all the events for the small pancreatic cancer monitoring cohort (Supplementary Figure 6C).

## Discussion

The results from the work demonstrate that a comprehensive whole-exome and transcriptome-wide liquid biopsy is a versatile solution to address the continuum of blood-based clinical applications, including early detection and potential screening for cancer, molecularly based cancer diagnosis, monitoring of MRD and recurrence in the early-stage curative approach setting, and precision selection of therapy for patients with advanced metastatic disease with targetable driver mutations.

Beginning with multi-cancer early detection, this broad genomic and technological approach leverages a plethora of somatic cfDNA features including base-pair alterations within their respective genomic contexts, copy number and aneuploidy, and fragment size dynamics. When these features were appropriately selected to maximize cancer specificity, we demonstrated the detection of malignancies at their earliest stages across various solid tumor types with a PPV higher than any other clinical platform to date. The clinical utility of this MCED approach cannot be understated given that cancers without a USPSTF screening modality account for ∼70% of all cancer deaths in the United States ^53^. A simple blood draw that can detect most early-stage incident cancers is poised to dramatically improve stage migration at diagnosis and the subsequent overall survival outcomes. Another vitally important aspect of our approach was the maintenance of a high specificity of 99.6%. False positives from the wide use of an MCED test can cause substantial undue burden for patients undergoing unnecessary scans and invasive biopsies and negative financial implications for the patient and health care system.

An inherent advantage of our approach is the measurement of the entire exome and transcriptome in circulation which provides more cancer features than hotspot approaches. The heterogeneity of cancer makes comprehensive identification of all hotspots challenging and thus existing methods that do not measure all potential alterations are inherently limited. Measuring hundreds of specific loci does not generate the same breadth of information as is achieved by measuring 49.7MB across the genome. Extrapolating the performance shown here to broader populations across various thresholds shows that analyzing all of the cell free DNA and RNA into circulation could have a significant benefit for patients (Supplementary Figures 7,8).

In the early stage/curative setting, numerous prior studies have demonstrated that the application of other currently existing liquid biopsy techniques for MRD and recurrence monitoring to detect micrometastatic disease after completion of definitive therapy delivered with curative intent has a stronger prognostic significance than any other clinical parameter, including stage, grade, tumor size, and nodal status ^54–57^. Furthermore, multiple lines of evidence in various cancers have shown that treatment escalation in MRD-positive patients may provide clinical benefit^58^. For example, recent data in early-stage colon cancer have shown that patients who are MRD-positive on liquid biopsy benefit from the use of adjuvant chemotherapy, whereas MRD-negative patients do not ^59^. Importantly, the approach employed to detect MRD does not rely on *a priori* genomic information from the primary tumor. This results in logistic efficiencies that require only a blood draw and avoids the need to procure and sequence tumor tissue. Furthermore, the use of a broad genomic approaches that sequences all exonic regions provides more “shots on goal” to identify cancer-derived features than smaller targeted approaches. A comparison of our approach with traditional targeted tumor-informed approaches will be the subject of future studies.

A limitation of our study is the limited sample size for certain cancer type and stage combinations, which may affect the generalizability of our findings. We are currently running a prospective study to more comprehensively assess ABCDai’s potential as an MCED.

In summary, the data presented here demonstrate a unified liquid biopsy platform that uses blood-based whole-exome and transcriptome sequencing coupled with artificial intelligence to address the important clinical needs in multi-cancer early detection, monitoring of MRD and recurrent cancers, and precision selection of molecularly targeted therapies.

## Data Availability

De-identified data is available upon request for academic or non-profit research purposes. To request access, please contact Caris legal at. Access is subject to review by the Caris legal team and approval will only be granted following execution of a formal agreement with acceptable terms. Requests will generally be processed within 6 months.

## Code Availability

Code used in this study is available upon request for academic or non-profit research purposes. To request access, please contact Caris legal at. Access is subject to review by the Caris legal team and approval will only be granted following execution of a formal agreement with acceptable terms. Requests will generally be processed within 6 months.

## Ethics Statement

Disclosure of Potential Conflicts of Interest: JA, VD, NP, SK, SA, JX, DAS, SS, RH, AS, JS, DDH, MO, MR, GWS and DS are employees of Caris Life Sciences. GP is a member of the board of directors of CLS. JLM, GDD, and AB serve on the scientific advisory board of Caris Life Sciences. All of the above have equity and/or equity options in Caris Life Sciences. EH, EL, SVL, and WSE have unpaid consultant/advisory board relationships with Caris Life Sciences. AFS serves on the consultant/advisory board, is on the speaker’s bureau, and has received travel funding from Caris Life Sciences.

SVL reports advisory role for Abbvie, Amgen, AstraZeneca, Boehringer Ingelheim, Bristol-Myers Squibb, Catalyst, Daiichi Sankyo, Eisai, Elevation Oncology, Genentech/Roche, Gilead, Guardant Health, Janssen, Jazz Pharmaceuticals, Merck, Merus, Mirati, Novartis, Pfizer, Regeneron, Sanofi, Takeda, and Turning Point Therapeutics; research grant (to institution) from Abbvie, Alkermes, Elevation Oncology, Ellipses, Genentech, Gilead, Merck, Merus, Nuvalent, RAPT, and Turning Point Therapeutics; and serving on a Data Safety Monitoring Board for Candel Therapeutics.YN reports advisory role from Guardant Health Pte Ltd., Natera,Inc., Roche Ltd., Seagen,Inc., Premo Partners, Inc., Daiichi Sankyo Co., Ltd., Takeda Pharmaceutical Co., Ltd., Exact Sciences Corporation, Gilead Sciences, Inc.; speakers’ bureau from Guardant Health Pte Ltd., MSD K.K., Eisai Co., Ltd., Zeria Pharmaceutical Co., Ltd., Miyarisan Pharmaceutical Co., Ltd., Merck Biopharma Co., Ltd., CareNet,Inc., Hisamitsu Pharmaceutical Co., Inc., Taiho Pharmaceutical Co., Ltd., Daiichi Sankyo Co., Ltd., Chugai Pharmaceutical Co., Ltd., Becton, Dickinson and Company, Guardant Health Japan Corp; research funding from Seagen,Inc., Genomedia Inc., Guardant Health AMEA, Inc., Guardant Health, Inc., Tempus Labs, Inc., Roche Diagnostics K.K., Daiichi Sankyo Co., Ltd., Chugai Pharmaceutical Co., Ltd. TY research funding from Amgen K.K., Chugai Pharmaceutical Co., Ltd., Daiichi Sankyo Co., Ltd., Eisai Co., Ltd., FALCO biosystems Ltd., Genomedia Inc., Molecular Health GmbH, MSD K.K., Nippon Boehringer Ingelheim Co., Ltd., Ono Pharmaceutical Co., Ltd., Pfizer Japan Inc., Roche Diagnostics K.K., Sanofi K.K., Sysmex Corp. and Taiho Pharmaceutical Co., Ltd.; honoraria for lectures from Chugai Pharmaceutical Co., Ltd., MSD K.K., Ono Pharmaceutical Co., Ltd., Bayer Yakuhin, Ltd., Merck Biopharma Co., Ltd. and Takeda Pharmaceutical Co., Ltd.; Consulting fees from Sumitomo Corp. GDD has received institutional support for oncology research studies to Dana-Farber Cancer Institute from Adaptimmune, Bayer, Novartis, PharmaMar, and Daiichi-Sankyo; he is also is a co-founder and consulting scientific advisory board member with minor equity holding in IDRx; a consultant/SAB member with minor equity holding in Erasca Pharmaceuticals, RELAY Therapeutics, Bessor Pharmaceuticals, CellCarta, Ikena Oncology, Kojin Therapeutics, Aadi Biosciences, Acrivon Therapeutics, Blueprint Medicines, Tessellate Bio, and Boundless Bio; he is also a scientific consultant for EMD-Serono/Merck KGaA, WCG/Arsenal Capital, and Minghui Pharmaceuticals.

