## Supplementary Material for "AI enabled exome and transcriptome liquid biopsy platform spanning the continuum of care in oncology"

### Validation of an AI-enabled exome/transcriptome liquid biopsy platform for early detection, MRD, disease monitoring, and therapy selection for solid tumors

#### Supplementary Methods

##### *Library Prep*

The cfTNA was extracted from plasma using a novel, high-throughput automated method, customized from the DSP Virus/Pathogen Midi kit (Qiagen, custom) and Hamilton Star liquid handler system. cfDNA/RNA was extracted from paired FFPE tissue using the manual FFPE DNA extraction kit (Qiagen) and RNeasy FFPE extraction kit (Qiagen) respectively, with the following modifications to preserve RNA molecules. First plasma was mixed with standard lysis buffer with high concentration of guanidinium salts, dithiothreitol (DTT), and carrier RNA to inhibit RNAases. Second, proteinase K was added to degrade protein RNAase molecules. Third SDS was added to standard binding buffer to lyse circulating microvesicles which were protecting RNA, and subsequent steps follow the standard protocol. All paired buffy coat samples had gDNA extracted using either the automated DSP DNA Midi kit (Qiagen, Cat# 937255) or the Mag-Bind Blood & Tissue DNA HDQ 96 Kit (Omega Biotek, Cat# M6399-01). Sequencing libraries were prepared using HyperPrep kits, HyperPure Beads, custom primer mixes and baits, (KAPA/Roche - Catalog No. 9983759001) and custom cDNA primers (IDT/GeneLink, AeG2A). Sequencing was performed on a NovaSeq System with NovaSeq 6000 S1 and S2 Reagent Kits (Illumina).

##### *NGS Output Processing*

Caris Assure is a hybrid assay where cell-free total nucleic acids (DNA and RNA) are sequenced together. Total FASTQ files are divided into DNA FASTQ and RNA FASTQ using the tag on the modified primer in addition to aligning the cf-TNA FastQ file to a Custom Genomic and Transcriptomic Reference. The reads in the BAM file that were aligned to the Transcript Reference and those with the tag were identified as RNA reads. These RNA reads and their mates were then written into RNA FastQ files and the remaining reads were written into DNA FastQ files. These DNA and RNA FastQ files were then aligned to the hg38 reference sequence using BWA-Mem (through sentieon-genomics bwa - 0.7.17-r1188). The resulting BAM files were analyzed to detect the anomalies present in the sample as described in the sections below.

*SNV/INDEL variant calling*

The BAM file for the sample was analyzed for the presence of SNV/Indel mutations using Mutect2 (through sentieon-genomics-202112.01 driver binary). The variants that were in the exclusion list of frequently detected false-positive variants, created during the initial validation phase of the pipeline, were removed. Only the pathogenic and likely pathogenic variants were reported. False positives were removed using the following thresholds:

- Minimum Base Quality (MBQ) of alternate (ALT) sequence >= 30
- Minimum Mapping Quality (MMQ) of ALT >= 20
- MBQ of ALT /MBQ of REF >= 0.5
- Allele Frequency (AF) > 0.001
- Median Position (MPOS) of variant in alternate reads >= 9
- Tumor LOD (TLOD) > 4, for lineage relevant variants, 6.3 otherwise.

*Variant Source*

The variants detected in the plasma were characterized by comparing the plasma and buffy coat sequencing results. A variant is characterized as being of tumor origin if the lower 95 CI of the plasma AF is greater than the upper bound of the 95 CI of the buffy coat AF. The variant is characterized as being of CH (clonal hematopoiesis) origin if the buffy coat AF is less than 20% and the 95 CI overlaps or is greater than the plasma AF. It is characterized as germline if the buffy coat AF is 30% or higher and the plasma AF is less than the buffy coat AF. The variant source is unknown if the buffy coat AF was between 20% and 30%. Variants detected in genes that are known to commonly harbor CH mutations are characterized as CH if the buffy coat AF is greater than the plasma AF.

*Copy Number Alteration for therapy selection*

The copy number alteration was measured using CNVKit version 0.9.10 (https://cnvkit.readthedocs.io/en/stable/)[1]. A gene is called amplified if the following criteria are met:

- lower bound of the gene segment confidence interval mean> 0.75
- segment weight > 100
- Weighted log2 ratio mean of the gene > 0.8
- standard deviation of all the genes on the sample < 1

*Machine Learning Methodologies for ABCDai-MCED*

A single ABCDai base model was trained on the entirety of the data from 507 patients with cancer and 506 normal donors. Performance was assessed on an independent set of 526 patients with cancer and 385 samples from normal donors.

*Machine Learning Methodologies for ABCDai-GPS*

ABCDai-GPS models were developed using 506 Normal samples and 660 stage I and II cancer samples. The samples were grouped into a normal category and 8 diagnostic pathways according to Supplementary Table 7. Multiclass modeling was performed to determine the correct diagnostic pathway. The performance was evaluated out of fold using 5-fold cross-validation.

*Machine Learning Methodologies for ABCDai-M&M*

A single ABCDai base model was trained on the entirety of the data from 1290 patients with cancer and 2149 normal donors. Performance was assessed on an independent set of 172 patients with known relapse status who had blood drawn after surgery. For patients in the MRD cohort, the first blood draw after surgery and before therapy was used and for the recurrence monitoring cohort, the last blood draw was used.

*MRD/Monitoring Survival Analysis*

Univariate analysis was conducted using Kaplan-Meier analysis, with significance determined using a logrank test. This was complemented by univariate analyses and controlled against common clinicopathological variables (age at collection, gender, BMI, and stage at collection) with multivariate hazard ratios obtained by fitting a Cox-proportional hazard model with a non-parametric Breslow baseline estimation. Significance was determined at p-values less than 0.05 for all analyses. For this analysis the limited set of Stage II/III labeled samples were conservatively treated as Stage III. Time-to-event analysis was performed using Python 3 (Python Software Foundation, https://www.python.org/) and the Lifelines (0.27.8) library [2].

MRD status was assessed at the initial blood-draw sample following surgery, prior to the initiation of adjuvant therapy. Monitoring status was evaluated at the final blood draw for patients who underwent at least one subsequent blood draw during adjuvant therapy. Both samples were run through ABCDai-M&M and High-risk status ("Relapse Predicted") was conferred by a positive model classification at the default probability threshold of 0.5.

*Statistical Analysis and Software*

An independent two-sample t-test was used to assess the significance of the mean differences for continuous variables with effect calculated with the Cohen D test. For determining significance between 3 of more groups, and when normality can be determined (though interpretation of the QQ plot), an Analysis of Variance (ANOVA) was performed followed by a post-hoc Tukey test to determine specific group-to-group differences. When normality could not be assumed, the Kruskal-Wallace test, followed by the post-hoc Dunn test, was used. For multiple group analysis effect was determined with the eta$^2$ test based on the H-statistic.

The Fisher's Exact test was used to assess the significance of differences in various performance metrics across models. Specifically, it compared true positive vs. false negative proportions to evaluate sensitivities, true positive vs. false positive proportions for PPV, true negative vs. false negative proportions for NPV, and true negative vs. false positive proportions for specificity. For Copyome distribution differences, across chromosome bands, the 2-sample Kolmogorov–Smirnov (KS) test was used. When appropriate, the Bonferroni correction was applied to control for multiple hypothesis testing. All statistical analysis was done with the python libraries statsmodels (0.14.4), scipy (1.13.0), and scikit_posthocs (0.9.1).

#### Ethics Statement

This study was conducted in accordance with the guidelines of the Declaration of Helsinki, Belmont report, and U.S. Common rule. In keeping with 45 CFR 46.101(b)(4), this study utilized retrospective, de-identified clinical data. Therefore, this study was considered IRB exempt and no patient consent was necessary from the subject. Approved protocol number WCG: TCBIO-001-0710.

2. Davidson-Pilon. lifelines: survival analysis in Python. https://doi.org/10.21105/joss.01317

#### Supplementary Tables and Table Legends

##### Supplementary Table 1. Sample Source Information

| Source | Number of Institutions | Consent | Samples: ABCDai_ M&M | ABCDai_ MCED | Normal |
|---|---|---|---|---|---|
| Caris_Biorepository | 80 | Patient Consent Obtained | 726 | 565 | 2149 |
| Caris_Clinical_Sample | 261 | Waiver Of Consent - WCG: TCBIO-001-0710 | 577 | - | - |
| Discovery_Life_Sciences_ Biorepository | Data Not Available | Patient Consent Obtained | - | 798 | - |
| Indivumed_Biorepository | 2 | Patient Consent Obtained | 172 | 176 | - |

##### Supplementary Table 2. Sample counts by Cancer Origin Tissue and stage

| Cancer Origin Tissue | Stage: 1 | 2 | 3 | 4 | Unknown |
|---|---|---|---|---|---|
| Bladder | 9 | 8 | 12 | 29 | 0 |
| Brain | 0 | 0 | 0 | 13 | 0 |
| Breast | 239 | 65 | 13 | 59 | 196 |
| Cancer of Unknown Primary | 0 | 0 | 0 | 27 | 0 |
| Cervical | 44 | 8 | 5 | 9 | 0 |
| Cholangiocarcinoma | 4 | 17 | 5 | 30 | 0 |
| Colorectal | 0 | 58 | 26 | 163 | 62 |
| Esophageal | 0 | 2 | 17 | 28 | 0 |
| Female Genital Tract Malignancy | 0 | 0 | 3 | 6 | 0 |
| GIST | 0 | 0 | 1 | 2 | 0 |
| Gastric | 5 | 24 | 25 | 18 | 0 |
| Head and Neck | 2 | 13 | 21 | 36 | 0 |
| Kidney Cancer | 163 | 15 | 6 | 5 | 0 |
| Liver | 2 | 5 | 2 | 4 | 0 |
| Lung NSCLC | 22 | 10 | 29 | 149 | 0 |
| Lung SCLC | 0 | 0 | 2 | 11 | 0 |
| Lymphoma | 5 | 3 | 12 | 4 | 0 |
| Melanoma | 0 | 9 | 1 | 14 | 0 |

| | | | | | |
|---|---|---|---|---|---|
| Neuroendocrine tumors | 1 | 0 | 2 | 10 | 0 |
| Other | 0 | 0 | 2 | 10 | 0 |
| Ovarian | 5 | 1 | 23 | 19 | 0 |
| Pancreatic | 7 | 18 | 2 | 49 | 0 |
| Prostate | 30 | 37 | 15 | 23 | 2 |
| Sarcoma | 2 | 2 | 7 | 3 | 0 |
| Small Intestine | 3 | 5 | 11 | 8 | 0 |
| Thymus | 2 | 0 | 0 | 1 | 0 |
| Thyroid | 18 | 1 | 1 | 2 | 0 |
| Urinary tract cancer | 3 | 1 | 0 | 1 | 0 |
| Uterus | 147 | 15 | 26 | 27 | 0 |
| Uveal Melanoma | 0 | 0 | 1 | 3 | 0 |

#### Supplementary Table 3. Lineage Performance by Stage

| Cancer Type | Stage | TP | FN | Sens (%) |
|---|---|---|---|---|
| ALL | 1 | 236 | 48 | 83.1% |
| Bladder | 1 | 1 | 0 | 100.0% |
| Breast | 1 | 4 | 2 | 66.7% |
| Cervical | 1 | 7 | 1 | 87.5% |
| Cholangiocarcinoma | 1 | 3 | 0 | 100.0% |
| Colorectal | 1 | 0 | 0 | 0.0% |
| Gastric | 1 | 3 | 1 | 75.0% |
| Head and Neck | 1 | 0 | 1 | 0.0% |
| Kidney Cancer | 1 | 110 | 16 | 87.3% |
| Liver | 1 | 0 | 1 | 0.0% |
| Lung NSCLC | 1 | 7 | 5 | 58.3% |
| Melanoma | 1 | 0 | 0 | 0.0% |
| Ovarian | 1 | 1 | 0 | 100.0% |
| Pancreatic | 1 | 0 | 0 | 0.0% |
| Prostate | 1 | 0 | 0 | 0.0% |
| Sarcoma | 1 | 2 | 0 | 100.0% |
| Small Intestine | 1 | 1 | 2 | 33.3% |
| Thymus | 1 | 0 | 2 | 0.0% |
| Thyroid | 1 | 7 | 9 | 43.8% |
| Urinary tract cancer | 1 | 1 | 1 | 50.0% |
| Uterus | 1 | 89 | 7 | 92.7% |
| ALL | 2 | 111 | 18 | 86.0% |

| | | | | |
|---|---|---|---|---|
| Bladder | 2 | 0 | 0 | 0.0% |
| Breast | 2 | 9 | 4 | 69.2% |
| Cervical | 2 | 2 | 0 | 100.0% |
| Cholangiocarcinoma | 2 | 14 | 0 | 100.0% |
| Colorectal | 2 | 23 | 2 | 92.0% |
| Gastric | 2 | 17 | 0 | 100.0% |
| Head and Neck | 2 | 5 | 1 | 83.3% |
| Kidney Cancer | 2 | 6 | 1 | 85.7% |
| Liver | 2 | 3 | 0 | 100.0% |
| Lung NSCLC | 2 | 1 | 1 | 50.0% |
| Melanoma | 2 | 4 | 2 | 66.7% |
| Ovarian | 2 | 1 | 0 | 100.0% |
| Pancreatic | 2 | 5 | 1 | 83.3% |
| Prostate | 2 | 5 | 3 | 62.5% |
| Sarcoma | 2 | 1 | 0 | 100.0% |
| Small Intestine | 2 | 5 | 0 | 100.0% |
| Thymus | 2 | 0 | 0 | 0.0% |
| Thyroid | 2 | 1 | 0 | 100.0% |
| Urinary tract cancer | 2 | 1 | 0 | 100.0% |
| Uterus | 2 | 8 | 3 | 72.7% |
| ALL | 3 | 76 | 14 | 84.4% |
| Bladder | 3 | 0 | 0 | 0.0% |
| Breast | 3 | 2 | 0 | 100.0% |
| Cervical | 3 | 1 | 0 | 100.0% |
| Cholangiocarcinoma | 3 | 1 | 0 | 100.0% |
| Colorectal | 3 | 12 | 0 | 100.0% |
| Gastric | 3 | 14 | 1 | 93.3% |
| Head and Neck | 3 | 12 | 0 | 100.0% |
| Kidney Cancer | 3 | 3 | 3 | 50.0% |
| Liver | 3 | 0 | 0 | 0.0% |
| Lung NSCLC | 3 | 2 | 0 | 100.0% |
| Melanoma | 3 | 0 | 0 | 0.0% |
| Ovarian | 3 | 2 | 0 | 100.0% |
| Pancreatic | 3 | 0 | 0 | 0.0% |
| Prostate | 3 | 11 | 2 | 84.6% |
| Sarcoma | 3 | 2 | 1 | 66.7% |
| Small Intestine | 3 | 9 | 2 | 81.8% |
| Thymus | 3 | 0 | 0 | 0.0% |
| Thyroid | 3 | 0 | 1 | 0.0% |

| | | | | |
|---|---|---|---|---|
| Urinary tract cancer | 3 | 0 | 0 | 0.0% |
| Uterus | 3 | 5 | 4 | 55.6% |
| ALL | 4 | 22 | 1 | 95.7% |
| Bladder | 4 | 1 | 0 | 100.0% |
| Breast | 4 | 0 | 0 | 0.0% |
| Cervical | 4 | 0 | 0 | 0.0% |
| Cholangiocarcinoma | 4 | 0 | 0 | 0.0% |
| Colorectal | 4 | 1 | 0 | 100.0% |
| Gastric | 4 | 4 | 0 | 100.0% |
| Head and Neck | 4 | 7 | 0 | 100.0% |
| Kidney Cancer | 4 | 0 | 0 | 0.0% |
| Liver | 4 | 0 | 0 | 0.0% |
| Lung NSCLC | 4 | 0 | 0 | 0.0% |
| Melanoma | 4 | 0 | 0 | 0.0% |
| Ovarian | 4 | 0 | 0 | 0.0% |
| Pancreatic | 4 | 4 | 0 | 100.0% |
| Prostate | 4 | 0 | 0 | 0.0% |
| Sarcoma | 4 | 0 | 0 | 0.0% |
| Small Intestine | 4 | 2 | 1 | 66.7% |
| Thymus | 4 | 0 | 0 | 0.0% |
| Thyroid | 4 | 0 | 0 | 0.0% |
| Urinary tract cancer | 4 | 1 | 0 | 100.0% |
| Uterus | 4 | 2 | 0 | 100.0% |

**Supplementary Table 4. Diagnostic Pathway Predictive Performance.**

| | Predicted | | | | | | | |
|---|---|---|---|---|---|---|---|---|
| Actual | CT_Abdomen _Chest | Colonos copy | Endosc opy | Liver_ US | Mammog raph with MRI | Neck _US | PS A | Pelvic _US |
| CT_Abdomen _Chest | 155 | 10 | 3 | 4 | 0 | 4 | 7 | 48 |
| Colonoscopy | 26 | 11 | 2 | 0 | 0 | 0 | 2 | 15 |
| Endoscopy | 20 | 2 | 6 | 4 | 0 | 0 | 0 | 4 |
| Liver_US | 8 | 2 | 0 | 14 | 0 | 0 | 0 | 3 |

| | | | | | | | | |
|---|---|---|---|---|---|---|---|---|
| Mammograph with MRI | 5 | 0 | 0 | 0 | 1 | 0 | 0 | 8 |
| Neck_US | 10 | 2 | 0 | 0 | 0 | 1 | 1 | 9 |
| PSA | 10 | 1 | 0 | 0 | 0 | 1 | 52 | 0 |
| Pelvic_US | 25 | 4 | 0 | 0 | 1 | 2 | 0 | 177 |

**Supplementary Table 5. Univariate and multivariate table comparing hazard ratios for the MRD validation cohort ('Model Prediction') vs. common clinicopathological variables.**

| Variables | Univariate Analysis | | | Multivariate Analysis | | |
|---|---|---|---|---|---|---|
| | Hazard Ratio | 95% Confidence Interval | p-value | Hazard Ratio | 95% Confidence Interval | p-value |
| Disease Free Survival | | | | | | |
| **Model Prediction** | | | | | | |
| Recurrence Free* | 1.00 | * | * | 1.00 | * | * |
| Recurrence | 33.40 | (4.13-270.25) | <0.005 | 116.56 | (2.69-5054.31) | 0.013 |
| **Patient Age at Diagnosis** | | | | | | |
| <50 | 0.00 | (0.0-inf) | 0.143 | 0.00 | (0.0-8.03) | 0.147 |
| 50-59* | 1.00 | * | * | 1.00 | * | * |
| 60-69 | 1.64 | (0.37-7.32) | 0.525 | 3.99 | (0.29-55.24) | 0.302 |
| >=70 | 0.72 | (0.12-4.31) | 0.704 | 0.11 | (0.0-3.29) | 0.204 |
| **Gender** | | | | | | |
| Male* | 1.00 | * | * | 1.00 | * | * |
| Female | 0.87 | (0.23-3.23) | 0.831 | 8.79 | (0.34-224.33) | 0.188 |
| **BMI Group** | | | | | | |
| Healthy Weight* | 1.00 | * | * | 1.00 | * | * |
| Overweight | 0.47 | (0.05-4.49) | 0.505 | 0.08 | (0.0-1.93) | 0.121 |
| Obese | 0.95 | (0.1-9.13) | 0.941 | 0.15 | (0.01-2.7) | 0.200 |
| **Stage** | | | | | | |
| 2* | 1.00 | * | * | 1.00 | * | * |
| 3 | 4.06 | (0.51-32.49) | 0.152 | 17.76 | (0.63-499.92) | 0.091 |

**Reference Class*

**Supplementary Table 6. Univariate and multivariate table comparing hazard ratios for the Monitoring validation cohort ('Model Prediction') vs. common clinicopathological variables.**

| Variables | Univariate Analysis | | | Multivariate Analysis | | |
|---|---|---|---|---|---|---|
| | Hazard Ratio | 95% Confidence Interval | p-value | Hazard Ratio | 95% Confidence Interval | p-value |
| Disease Free Survival | | | | | | |
| **Model Prediction** | | | | | | |
| Recurrence Free* | 1.00 | * | * | 1.00 | * | * |
| Recurrence | 4.40 | (1.34-14.45) | 0.008 | 6.13 | (1.3-29.02) | 0.022 |
| **Patient Age at Diagnosis** | | | | | | |
| <50 | 0.00 | (0.0-inf) | 0.158 | 0.03 | (0.0-60.6) | 0.365 |
| 50-59* | 1.00 | * | * | 1.00 | * | * |
| 60-69 | 1.99 | (0.39-10.26) | 0.406 | 2.31 | (0.39-13.46) | 0.353 |
| >=70 | 1.25 | (0.23-6.83) | 0.812 | 0.61 | (0.09-4.17) | 0.617 |
| **Gender** | | | | | | |
| Male* | 1.00 | * | * | 1.00 | * | * |
| Female | 0.85 | (0.18-3.96) | 0.841 | 0.54 | (0.09-3.1) | 0.487 |
| **BMI Group** | | | | | | |
| Healthy Weight* | 1.00 | * | * | 1.00 | * | * |
| Overweight | 0.42 | (0.09-2.0) | 0.260 | 0.17 | (0.03-1.02) | 0.052 |
| Obese | 0.24 | (0.03-1.98) | 0.149 | 0.09 | (0.01-1.01) | 0.051 |
| **Stage** | | | | | | |
| 2* | 1.00 | * | * | 1.00 | * | * |
| 3 | 2.07 | (0.55-7.82) | 0.271 | 2.21 | (0.52-9.35) | 0.283 |

**Reference Class*

#### Supplementary Table 7. Proposed Diagnostic Pathways

| Diagnostic Pathway | Cancer Types |
|---|---|
| Colonoscopy | Colorectal |
| CT_Abdomen_Chest | Pancreatic, Kidney Cancer, Bladder, Urinary tract cancer, Lung NSCLC, Lung SCLC |
| Endoscopy | Esophageal, GIST, Gastric, Small Intestine |
| Liver_US | Cholangiocarcinoma, Liver |
| Mammograph with MRI | Breast |
| Neck_US | Head and Neck, Thyroid |
| Pelvic_US | Uterus, Cervical, Ovarian, Female Genital Tract Malignancy |
| PSA | Prostate |

#### Supplementary Figures and Figure Legends

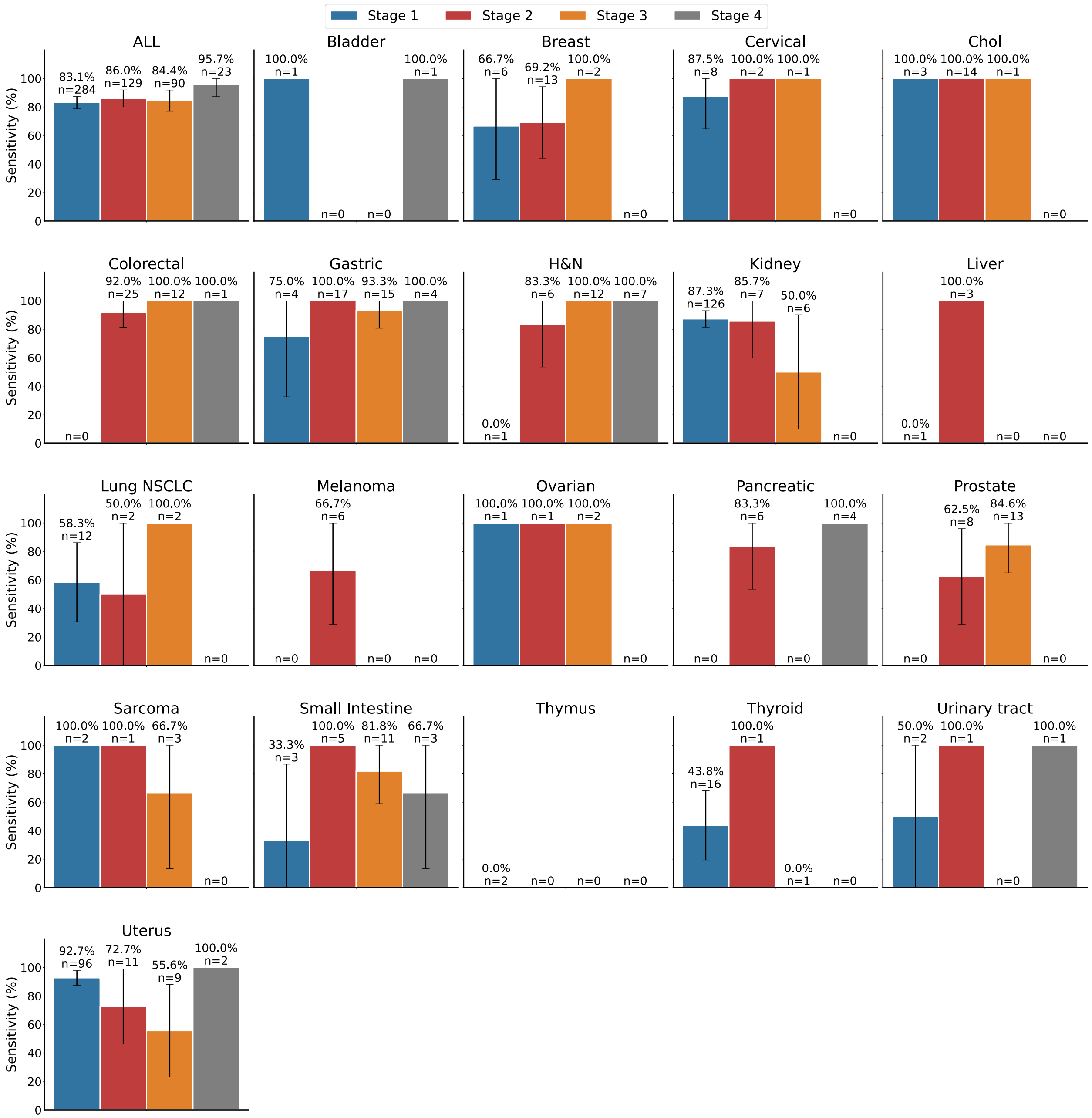


**Supplementary Figure 1.** Sensitivity of ABCDai by Tumor Type and Stage. Bar plots represent the ABCDai-MCED sensitivity on an independent data set.

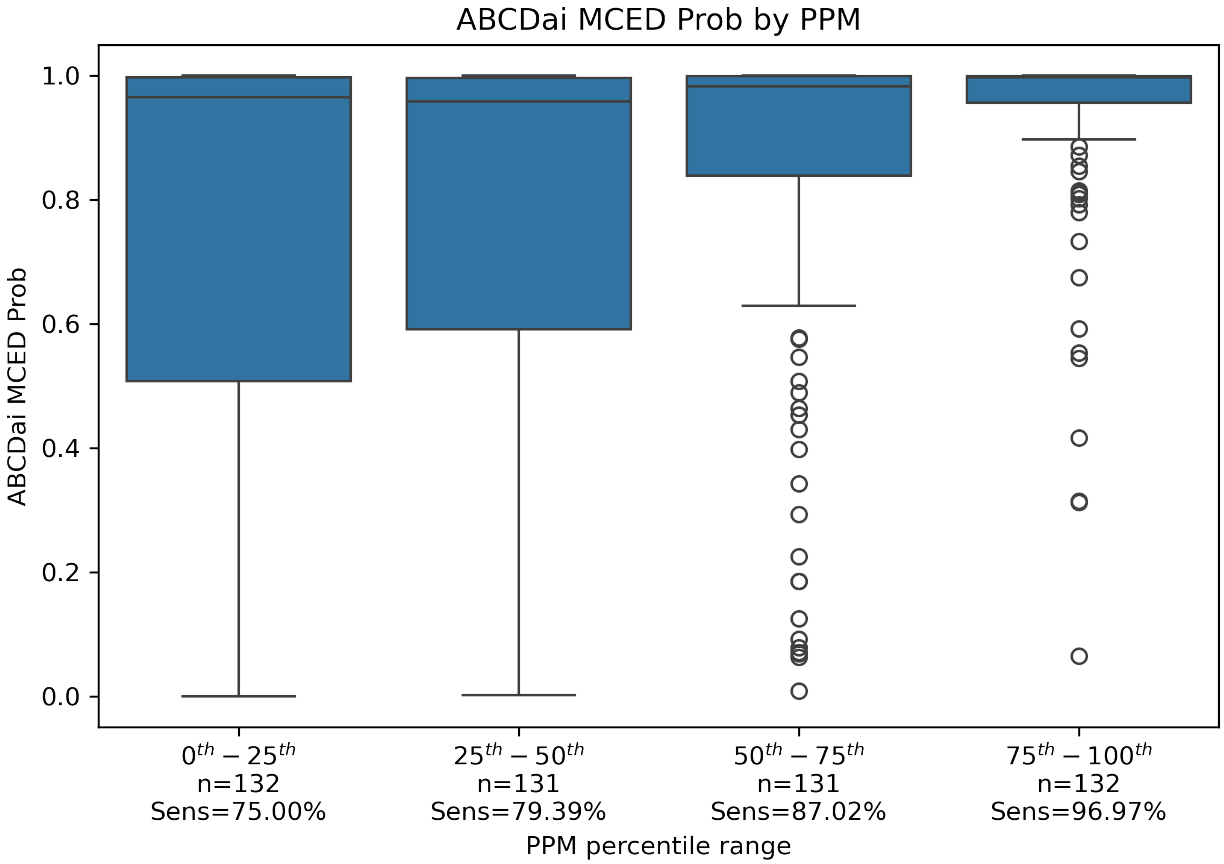


**Supplementary Figure 2.** ABCDai MCED Prob by PPM shows the distribution of ABCDai probability for the 526 samples in MCED Independent validation cancer samples, grouped by PPM Percentile range. The values for the bounds of the percentile ranges shown are: $0^{th}$: 0.00, $25^{th}$: 0.30, $50^{th}$: 0.52, $75^{th}$: 0.95, $100^{th}$: 8.77. The first 3 groups include the lower bound but not the upper bound. The last group includes both bounds, $75^{th}$ and $100^{th}$ percentile. The median probability for the groups is: $0^{th}$ - $25^{th}$: 0.965, $25^{th}$ - $50^{th}$: 0.959, $50^{th}$ – $75^{th}$: 0.983, $75^{th}$ – $100^{th}$: 0.996.

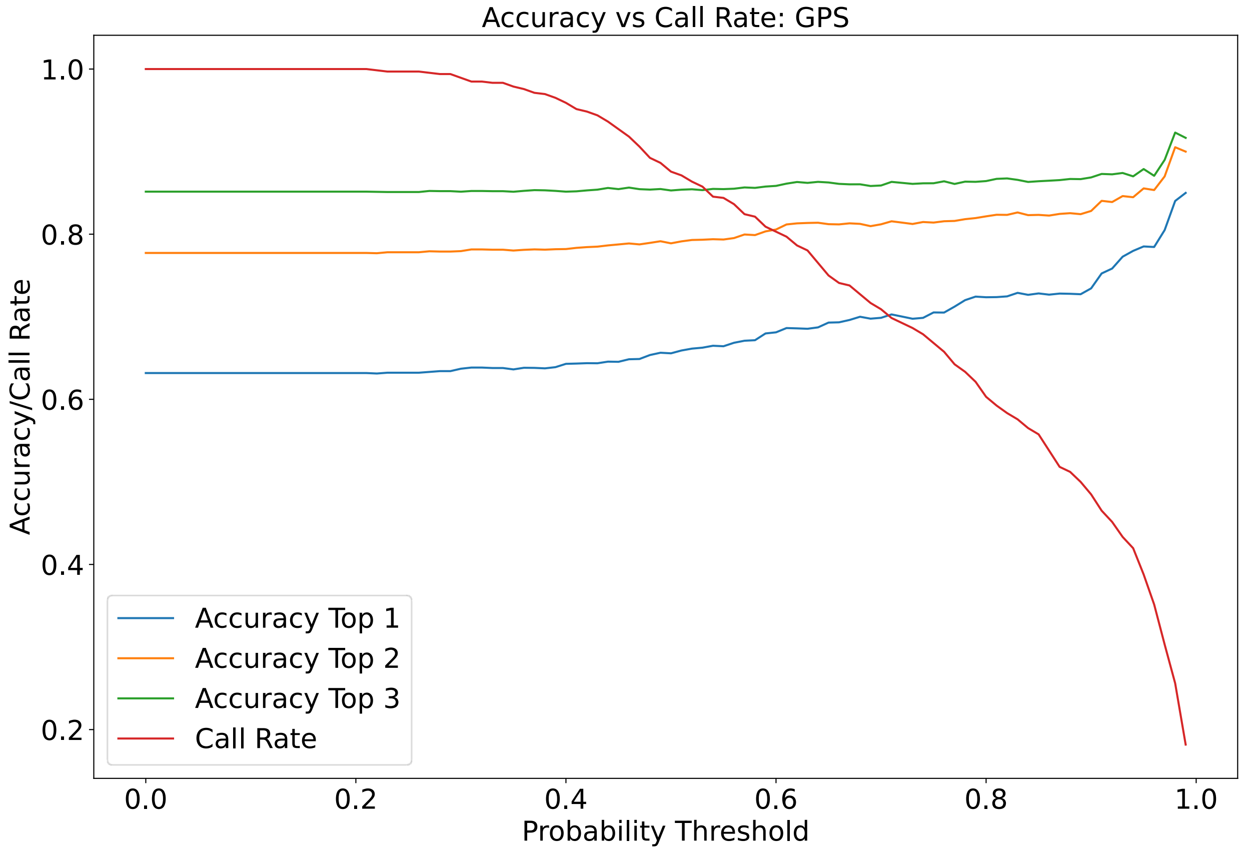


**Supplementary Figure 3.** Diagnostic Pathway Accuracy by Call Rate for Stage I Cancer

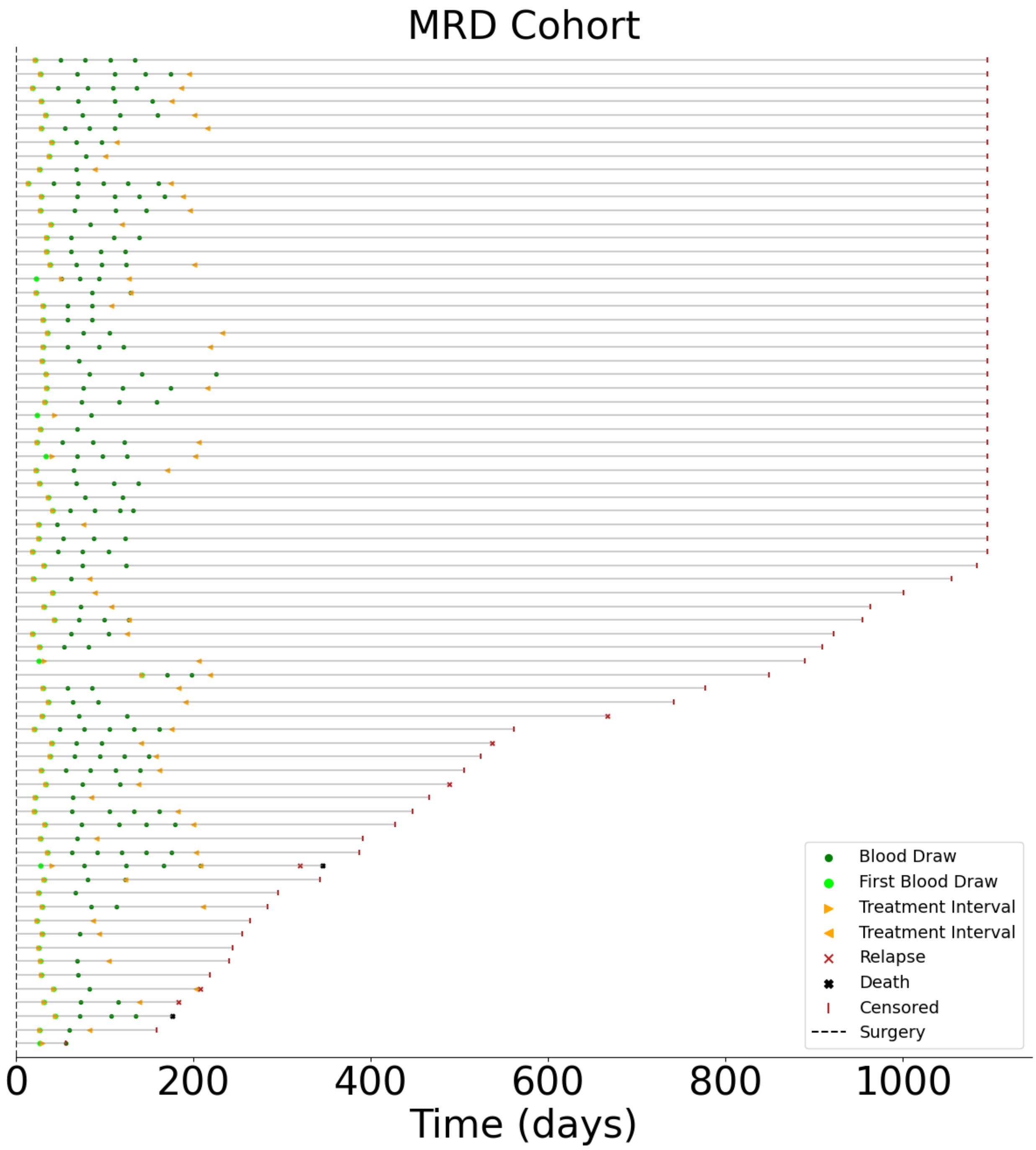


**Supplementary Figure 4.** Patient Treatment and Event Timelines for the MRD Cohort: An overview depicting the progression of the cohorts. Highlighted intervals include subsequent treatment phases and corresponding blood draws.

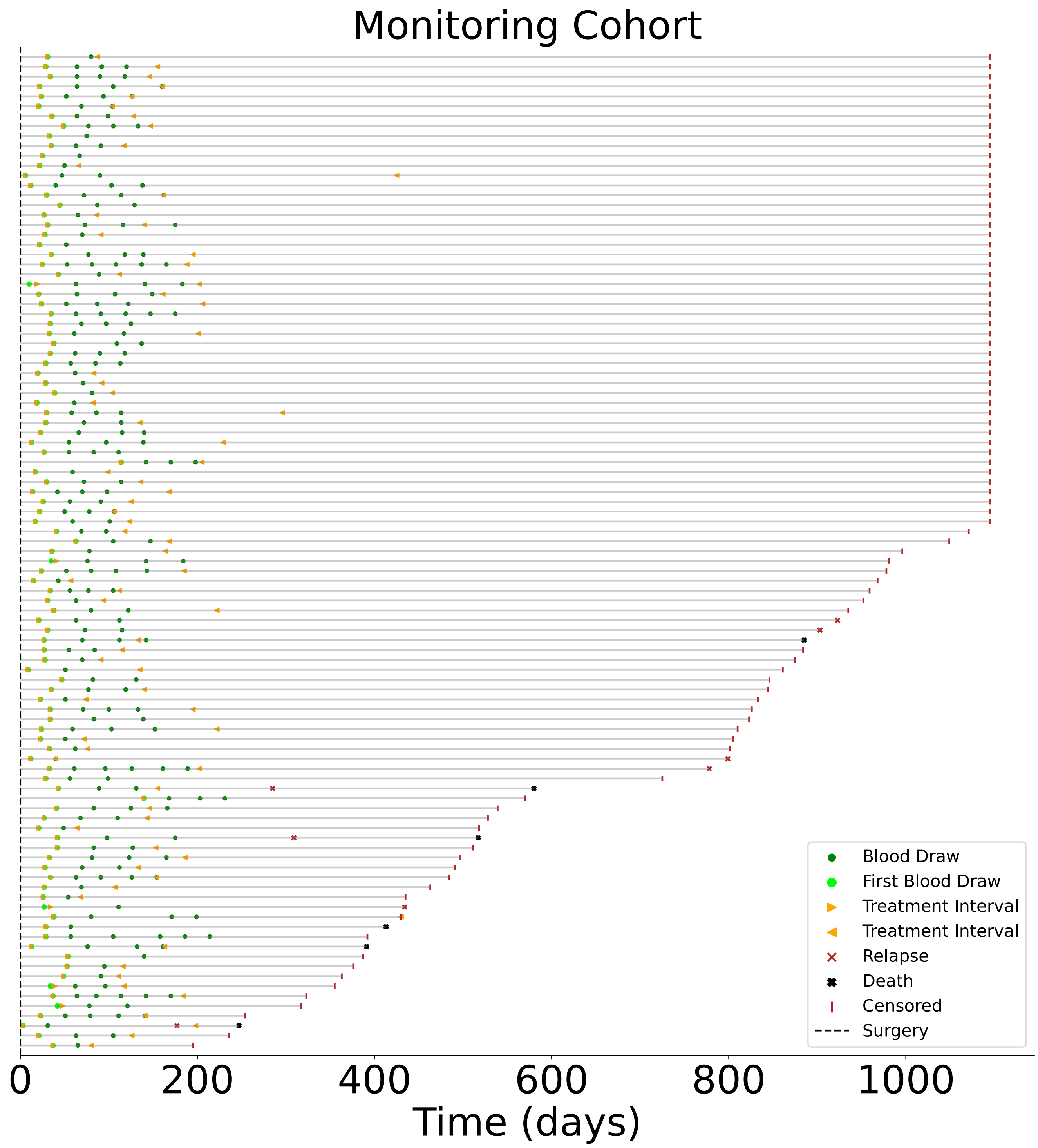


**Supplementary Figure 5.** Patient Treatment and Event Timelines for the Monitoring Cohort: An overview depicting the progression of the cohorts. Highlighted intervals include subsequent treatment phases and corresponding blood draws.

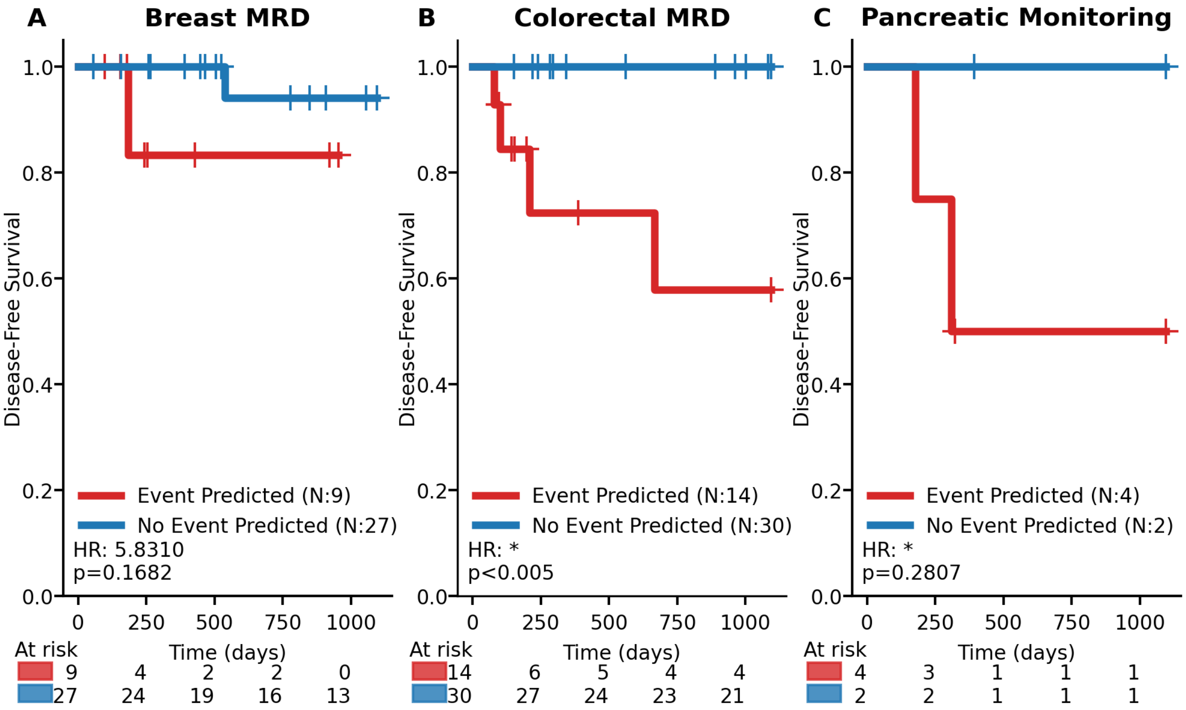


**Supplementary Figure 6.** ABCDai-M&M DFS performance on specific cancer types in the independent MRD and Monitoring Validation Cohorts. A) Kaplan-Meier survival curves for ABCDai-M&M applied on Breast MRD samples in the independent validation using the default probability threshold of 0.5. B) Kaplan-Meier survival curves for ABCDai-M&M applied on the Colorectal MRD samples in the independent validation using the default probability threshold of 0.5. C) Kaplan-Meier survival curves for ABCDai-M&M applied on the Pancreatic recurrence monitoring samples in the independent validation using the default probability threshold of 0.5. Univariate hazard ratios are derived from Cox Proportional Hazards models, with significance assessed via log-rank tests. Events are defined as relapse or death, with time measured from the date of surgery for MRD and from therapy initiation for monitoring. (*) indicates that the hazard ratio can't be computed due to a lack of events in the 'No Event Predicted' arm.

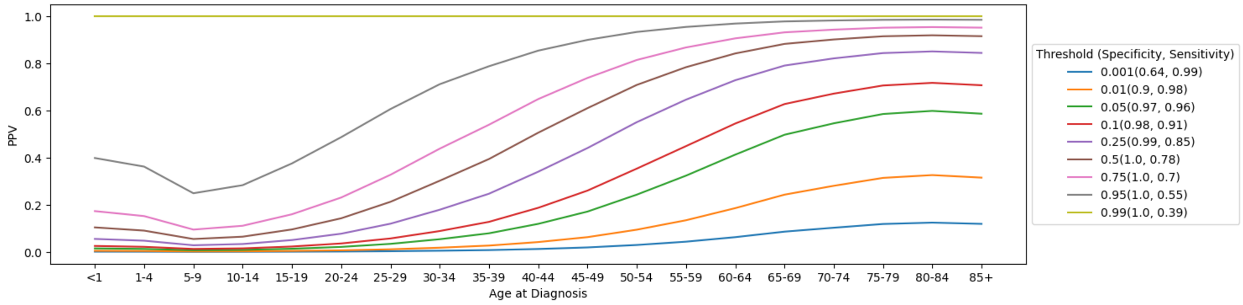


**Supplementary Figure 7.** ABCDai- MCED performance extrapolated to the general population as a function of probability threshold and age. Data from SEER was used to compute a weighted sensitivity adjusting for the prevalence of each cancer type.

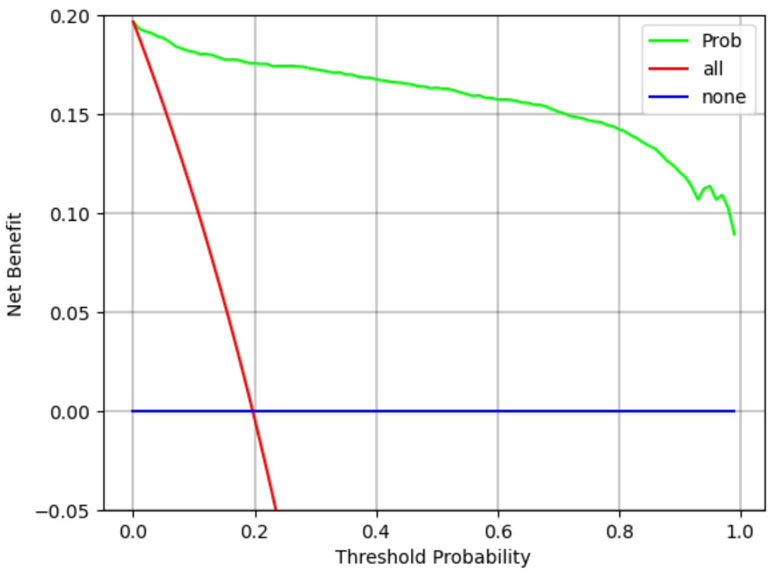


**Supplementary Figure 8.** ABCDai- Decision curve analysis shows that the test performance would deliver significant benefit.
